# A recessive *HNF1A* p.A251T variant causes monogenic diabetes by altering islet cell development, insulin secretion and intercellular connectivity

**DOI:** 10.1101/2024.12.10.24318788

**Authors:** Ines Cherkaoui, Marie Gasser, Qian Du, Dieter M. Egli, Camille Dion, Harry G. Leitch, Giada Ostinelli, Dilshad Sachedina, Shivani Misra, Guy A. Rutter

**Affiliations:** Centre de Recherche du CHUM, and Faculty of Medicine, University of Montreal, Montréal, QC, Canada; Department of Metabolism, Digestion and Reproduction, Imperial College London, UK; Departments of Pediatrics, Naomi Berrie Diabetes Center, Irving Medical Center, Columbia University, New York, USA; MRC Laboratory of Medical Sciences, West London, UK; UCL Great Ormond Street Institute of Child Health, Genetics & Genomic Medicine Department, London, UK and North East Thames Regional Genetic Service, Great Ormond Street Hospital for Children NHS Foundation Trust, London, UK; Department of Medicine & Integrated Care, Imperial College Healthcare NHS Trust, London, UK; Department of Dermatology, Imperial College Healthcare NHS Trust, London, UK; Lee Kong Chian Imperial Medical School, Nanyang Technological University, Singapore; Research Institute of the McGill University Health Sciences Center, Montreal, QC, Canada

**Author notes:** Address correspondence to: Professor Guy A. Rutter;<u>; +1</u> Or Dr Shivani Misra. Other author email addresses*: Ines Cherkaoui, Marie Gasser, Qian Du, Dieter Egli, Camille Dion, Harry Leitch, Giada Ostinelli, Dilshad Sachedina.

## Abstract

Monogenic diabetes results from single-gene defects that impair pancreatic beta-cell function, with mutations in the *HNF1A* gene being a common cause. While most *HNF1A* mutations exhibit dominant inheritance, the p.A251T (c.751G>A) variant demonstrates recessive transmission. To define its pathogenic role, induced pluripotent stem cells (iPSCs) derived from a patient homozygous for *HNF1A* p.A251T, and CRISPR/Cas9-edited human embryonic stem cells carrying the hemizygous variant, were differentiated into pancreatic islet-like cells. Islet-like clusters carrying the p.A251T variant showed impaired differentiation into insulin-producing cells, while alpha-cell formation was increased. Correspondingly, transcriptomic analysis revealed decreases in beta- and increases in alpha cell-enriched genes. Functional studies in islet-like clusters demonstrated lowered glucose-stimulated insulin secretion and diminished intracellular Ca^2+^ responses, including altered beta-cell connectivity. Responses to sulphonylureas and to glucagon-like peptide-1 receptor agonist were retained. These results indicate that the *HNF1A* p.A251T variant impairs beta-cell lineage specification and function. Our findings establish patient-derived and genome-edited stem cell models as a translational platform to assess the pathogenicity of rare *HNF1A* variants and to guide precision therapy for monogenic diabetes. We also define intercellular connectivity as a partially-preserved feature of stem cell-derived human beta-like cells and show that this is affected in a model of monogenic diabetes.

**Highlights:**

- Patient-derived and CRISPR/Cas9-edited human stem cells were used to model the rare recessive *HNF1A* p.A251T variant.
- The *HNF1A* p.A251T variant impairs pancreatic islet differentiation, reducing mature beta cells and increasing the alpha-like cell population.
- The variant compromises glucose-stimulated insulin secretion, which is partially rescued by sulphonylureas, and intercellular connectivity as assessed by Ca^2+^ imaging.
- This study demonstrates that patient-derived stem cell models can assess *HNF1A* variant pathogenicity and supports precision therapy for monogenic diabetes.

Graphical abstract.
Mechanistic impact of the *HNF1A* p.A251T variant on beta-cells identity and function. The *HNF1A* p.A251T mutation leads to multiple β-cell defects, including reduced glucose-stimulated insulin secretion and diminished intracellular Ca² response to high glucose. Additionally, there is decreased expression of β-cell commitment markers, suggesting impaired differentiation, and an increase in alpha-like cell populations. Together, these changes result in mildly disrupted β-cell identity and function, indicating partial β-cell functional insufficiency.

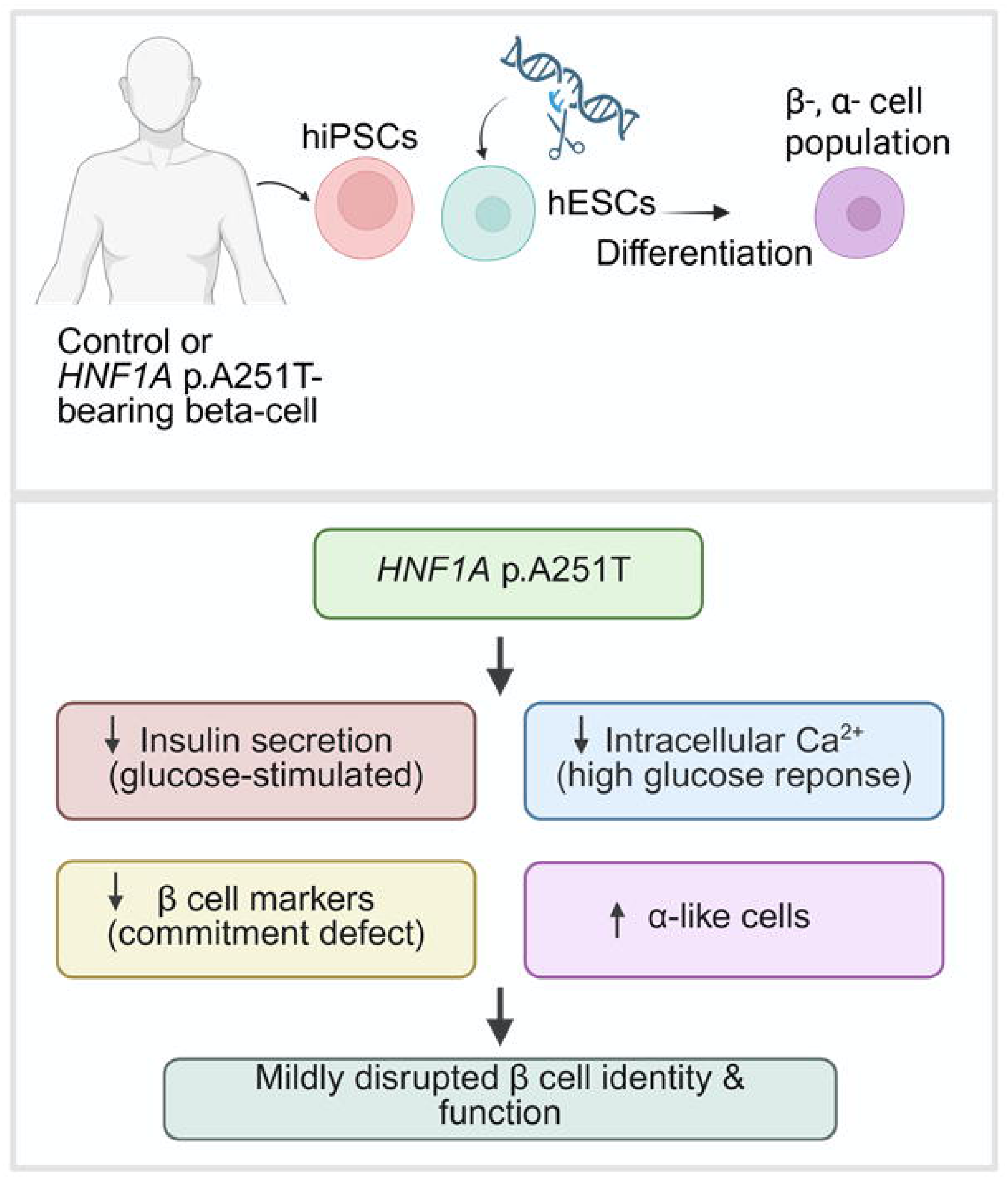

## Introduction

Monogenic diabetes represents a rare but clinically important subset of diabetes cases caused by mutations in single genes that impair pancreatic beta-cell function^1^. Among these, hepatocyte nuclear factor 1-alpha maturity-onset diabetes of the young (HNF1A-MODY) is the most common^2^. Accurate molecular diagnosis is essential, as individuals with HNF1A-MODY often respond well to sulfonylureas instead of insulin^3^, thereby avoiding the need for multiple daily injections and intensive glucose monitoring.

The use of next-generation sequencing has greatly improved the detection of variants in genes associated with monogenic diabetes. However, the increasing identification of variants of uncertain significance (VUS) presents a diagnostic challenge, as their clinical relevance and functional impact are unclear ^4,5^. Identifying the optimal treatment may require a stepwise, experience-guided evaluation of therapeutic alternatives.

In the present study, we investigated the pathogenicity of a rare *HNF1A* missense variant, p.A251T (c.751G>A), previously identified in a family with early-onset diabetes from the “MY DIABETES” study (ClinicalTrials.gov, NCT02082132)^6^. The proband and siblings carrying this variant in the homozygous state demonstrated sustained improvement in glycemic control following transition from insulin to low-dose sulfonylurea therapy, consistent with the HNF1A-MODY phenotype^3,6^. Our previous studies^6^ involved functional explorations using the overexpressed proteins in immortalized non-human beta cells. Here, we use patient-derived and CRISPR/Cas9-edited stem cell models to investigate how the *HNF1A* p.A251T variant affects pancreatic islet development and beta-cell function.

## Methods

### Human biopsies and ethical approval

The *Studying Rare Diabetes Using Induced Pluripotent Stem Cells (DIPS)* study, investigating rare forms of diabetes using patient-derived induced pluripotent stem cells, was approved by the Westminster Research Ethics Committee (Ref: 20/LO/1071). Skin biopsies were obtained from a participant homozygous for the *HNF1A* p.A251T variant after obtaining written informed consent in accordance with the Declaration of Helsinki.

### Plasmid construction and site-directed mutagenesis

Constructs were provided by the Oxford Centre for Diabetes Endocrinology and Metabolism^7^. A plasmid encoding human pancreatic *HNF1A wild-type* (WT) cDNA (NM_000545.8) in a pcDNA3.1 backbone was used for all functional assays. The *HNF1A* p.A251T (c.751G>A) variant was generated using the Q5 Site-Directed Mutagenesis Kit (New England BioLabs, USA). All constructs were sequenced (Genewiz, USA); primer sequences in Supplementary Table 1. Reporter assays employed pGL3 vectors (Promega, USA) containing HNF1A-binding sites upstream of either the rat albumin (pGL3-RA) or HNF4A P2 (pGL3-P2) promoter driving Firefly luciferase.

### Cell culture, transfection, transactivation and DNA binding assays

Functional assays were performed in HeLa cells^8^ as previously described^6,9^. Firefly and Renilla luciferase activities were measured using the Dual-Luciferase Reporter Assay System (Promega, USA) on a GloMax plate reader (Promega, USA). DNA-protein interactions were quantified using a colorimetric DNA-protein binding assay kit (Abcam, United Kingdom) according to the manufacturer’s instructions. Bound complexes were detected with an anti-HNF1A antibody (Santa Cruz Biotechnology, 1:1000, USA) and HRP-conjugated secondary antibody (GE Healthcare, 1:1000, USA) (Antibody details in Supplementary Table 2). Positive controls for HNF1A impaired protein function included the MODY-causing mutations p.P112L^10^, p.R203H^5^, and p.T260M^11^; and the p.E508K^12^ and p.A98V^13^ variants associated with an increased risk of T2D provided by the Oxford Centre for Diabetes Endocrinology and Metabolism^7^.

### Subcellular localization

INS1(832/3) cells were transiently transfected for 24h with WT or other variant HNF1A constructs used as controls. Subcellular localization was determined by immunofluorescence using an anti–6×His monoclonal antibody (ThermoFisher, USA, 1:1000) and Alexa Fluor 488–conjugated secondary antibody (ThermoFisher, USA, 1:1000).

### Skin punch biopsy and fibroblast culture

Using established protocols, epidermal biopsies on the arm (4 mm diameter) were obtained ^14^, cut into smaller sections and cultured at 37 °C with 5% CO_2_ in Dulbeccòs modified Eaglès medium (DMEM, ThermoFisher, USA) supplemented with 15% FBS (ThermoFisher, USA) and Antibiotic-Antimycotic cocktail (Gibco ThermoFisher, USA).

### Generation of iPSCs from Primary Fibroblasts

Human dermal fibroblasts (3 × 10 cells), at 80–90% confluence, were electroporated with episomal plasmids encoding *OCT4*, *SOX2*, *KLF4*, *L-MYC*, and *LIN28* (pCXLE-hOCT3/4-shp53-F, pCXLE-hSK, and pCXLE-hUL; Addgene plasmids #27077, #27078, and #27080, respectively). Reprogramming and culture conditions followed the manufacturer’s guidelines (ThermoFisher, USA, *Reprogramming Fibroblasts with Episomal Vectors*, Pub. No. MAN00077034).

### CRISPR/Cas9 genome editing of hESCs

*INS*(GFP/w) Human Embryonic Stem Cells (hESCs MEL1^15^; 2 × 10) were pre-treated with StemFlex medium containing 10 µM ROCK inhibitor (Y-27632) and electroporated with Alt-R CRISPR-Cas9 ribonucleoprotein (RNP) complex (Integrated DNA Technologies, USA) containing synthetic sgRNA, recombinant Cas9, and a single-stranded oligodeoxynucleotide (ssODN) template encoding the *HNF1A* p.A251T variant using a BTX Gemini electroporator (250 V, 5 μs, single square-wave pulse). GFP cells were FACS-sorted 48 h post-electroporation. Clones were expanded and genotyped by PCR, Sanger sequencing, and Amplicon-EZ (Genewiz, USA). HNF1A knockout (KO) cell lines used as controls were described previously ^16^.

### Directed differentiation to pancreatic lineages

Human pluripotent stem cells were differentiated into beta-like cells using a stepwise protocol adapted from published methods^17–19^ and harvested at stage 6 (“beta-like cells”).

### Immunofluorescence Staining

Cryosections of 4% PFA (Paraformaldehyde)-fixed islet-like clusters (day 25) were prepared and imaged as per^17,18^.

### Glucose-Stimulated Insulin Secretion (GSIS)

Assays were performed in Krebs-based medium as described^17^. Insulin was measured with an ultra-sensitive homogenous time-resolved fluorescence (HTRF) assay (Cisbio, France), according to the manufacturer’s instructions.

### Intracellular free Calcium (Ca^2+^) imaging

Islet-like clusters (stage 6) were incubated with Cal-590 AM (5 µM; AAT Bioquest, USA) in glucose/glutamine-free DMEM (ThermoFisher, USA), containing 3.3mM glucose, for 1 h at 37°C, then imaged on a Zeiss Axiovert spinning disk microscope. Baseline (3min, 3.3mM glucose) was followed by a stepwise glucose ramp (11 mM to 16.7 mM, 15 min each), ending with 20mM KCl (positive control). Ca^2+^ traces of insulin were analyzed for connectivity as previously described^20^, with optimization for islet-like clusters: a signal threshold of ΔF/F > 2% was used.

### RNA Extraction, RT-qPCR

Total RNA was extracted from day 27 beta-like clusters using the RNeasy Mini Kit (Qiagen, Germany). cDNA was synthesized from 10 ng RNA using the High-Capacity cDNA Kit (Thermo Fisher, USA). RT-qPCR was performed with PowerUp™ SYBR™ Green with standard cycling. Expression was normalized to ACTB and analyzed via the 2^−ΔΔCt^ method. Primer sequences are in Supplementary Table 1.

### RNA Sequencing

Raw FASTQ reads were processed using GenPipes v6.0.0^21^ quantified with kallisto v0.50.0^22^ against the GRCh38 Ensembl v90 genome assembly. Gene-level counts were derived from transcript abundance estimates. Differential expression analysis was performed with DESeq2 v1.48.1^23^, adjusting for batch effects. Gene set enrichment was conducted using fgsea v1.34.2^24^, and GO/pathway enrichment with gProfiler2 v0.2.3^25^. Analyses were run in R v4.5.0 under Rocky Linux 8.10.

### Flow cytometry

Endocrine cell clusters were dissociated into single cells using TrypLE Express (ThermoFisher, USA). Cells were fixed and permeabilized using the FIX & PERM Cell Fixation and Permeabilization Kit (ThermoFisher, USA) according to the manufacturer’s instructions. Samples were stained with an anti-insulin, anti-glucagon antibodies (details in Supplementary Table S2). Data were acquired using an Aurora CS flow cytometer (Cytek Biosciences) and analyzed with FlowJo software (BD Biosciences).

### Statistical analysis

All statistical analyses were performed using GraphPad Prism version 10 (San Diego, USA) and MATLAB R2022b (Natick, USA). Data distribution was assessed using the Shapiro–Wilk test^26^. A *P* value < 0.05 was considered statistically significant.

## Results

### Modest functional impairment of the HNF1A p.A251T variant assessed by overexpression in immortalized cell lines

The *HNF1A* p.A251T variant, generated by site-directed mutagenesis, was validated by Sanger sequencing (c.751G>A; Supplementary Figure S1). Alanine 251 lies within the DNA-binding domain of HNF1A (Figure 1A), which regulates transcription through nuclear localization and target gene interaction. Extending previous studies^6^ we found that, compared to WT, the p.A251T variant showed reduced nuclear localization (WT: 87.7 ±9.0 %, A251T: 80.6% ±11.2, p = 0.007; Figure 1B, 1D). Transactivation activity was largely preserved (A251T: 86.2±29.3% of WT; p=0.10; Figure 1C) and notably higher than pathogenic MODY variants p.P112L (30.3 ± 8.3%) and T260M (34.0 ± 17.4%) (p < 0.05; Figure 1F). The DNA-binding capacity of the A251T protein tended to be lower than WT (62.3±0.2 % of WT; p=0.12; Figure 1E). These results thus suggest that p.A251T variant modestly impairs nuclear localization but retains near-normal transactivation and DNA-binding activity.

**Figure 1.**
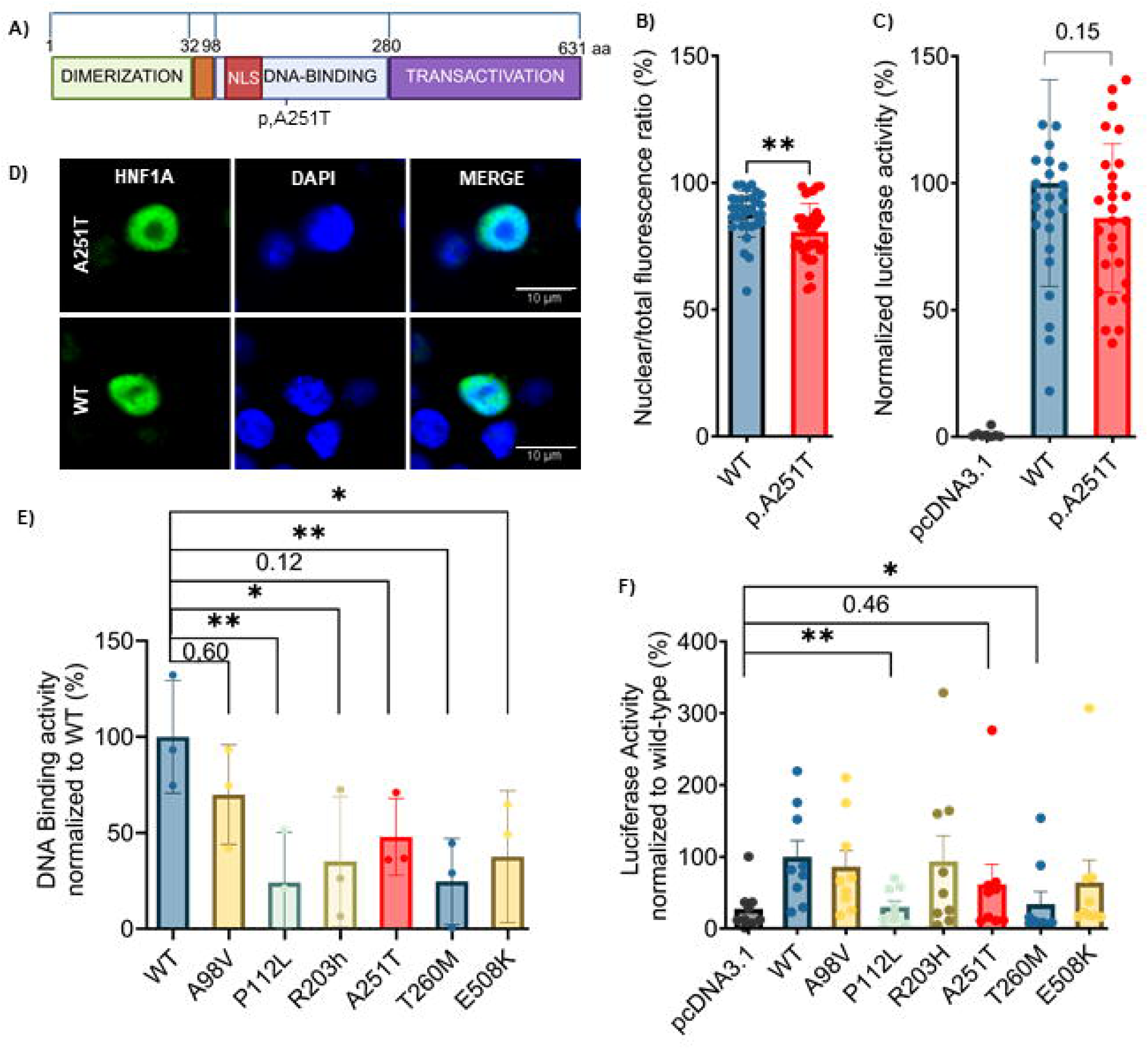
Functional characterization of *HNF1A* p.A251T variant in overexpression systems. (A) Schematic representation of HNF1A protein domains: dimerization (residues 1–32), DNA-binding (98–280), and transactivation (281–631). The *HNF1A* p.A251T variant is located within the DNA-binding domain. (B) Quantification of the nuclear-to-total fluorescence signal ratio in INS-1 832/3 cells transiently expressing 6×His-tagged WT or *HNF1A* p.A251T. (C) Transactivation activity in HeLa cells co-transfected with WT, p.A251T, or empty pcDNA3.1 vector, together with the pGL3-RA reporter (rat albumin promoter with an HNF1A-binding site) and pRL-SV40 Renilla luciferase control. Firefly luciferase activity is expressed as a percentage of WT activity and normalized to empty vector baseline. Each point represents the mean of nine technical replicates from three independent experiments. (D) Representative confocal images (×40 oil immersion; Nikon Eclipse T-I microscope) of INS-1 832/3 cells showing HNF1A localization (green, Alexa Fluor 488) and nuclei (blue, DAPI). (E) DNA-binding activity measured in HeLa cells using a biotin-labeled HNF1A consensus oligonucleotide and a colorimetric DNA-protein binding assay. Values are normalized to WT and corrected for empty vector baseline. (F) Transactivation activity of known pathogenic *HNF1A* variants (P112L, R203H, T260M) used as positive controls for loss-of-function or variant associated with a significantly higher risk for developing T2D (A98V, E508K) (n = 27); Significance *p ≤ 0.05; non-significant trends are indicated with exact p-values, not indicated when p>0.5.

### HNF1A p.A251T-bearing human stem cells display impaired beta-cell but enhanced alpha cell formation

To model dosage-dependent effects of *HNF1A* loss in a controlled genetic background, we used CRISPR/Cas9-mediated gene editing in *INS*(GFP/w) hESCs^15^. A heterozygous *HNF1A*^+/–^ line (p.Thr251fs333) and a hemizygous *HNF1A*^−/A251T^ line (p.A251T + p.Thr251fs333) were generated and validated by Sanger sequencing (Figure S2A–D). Western (immuno-) blotting revealed reduced HNF1A protein in all edited lines vs. WT cells (e.g. +/–: 28.5%; A251T/–: 24.4%; all p < 0.0001; Figure S2E–F). The *HNF1A*^−/–^hESC line was obtained from a previously validated model (Bryan J. González et al., 2022^16^).

To complement the isogenic model, dermal fibroblasts from a proband homozygous for the *HNF1A* p.A251T variant were reprogrammed into iPSCs (Figure S3A). The resulting colonies showed typical pluripotent morphology (Figure S3B), retained the homozygous p.A251T variant confirmed by Sanger sequencing (Figure S3C), and expressed nuclear OCT4 in 98.9 ± 0.6% of cells (n=17; Figure S3D, F), with no expression in the parental fibroblasts (p < 0.0001). All clones displayed normal karyotypes (Figure S3E) and robust alkaline phosphatase activity (Figure S3G).

All hPSC lines were differentiated into pancreatic islet-like clusters using a stepwise protocol^17–19^, with aphidicolin added at stage 5 to enhance β-cell maturation (Figure S4). Immunostaining revealed a significant reduction in the proportion of insulin-positive cells (INS) within islet-like clusters derived from *HNF1A*^A251T/A251T^ iPSCs compared to control iPSCs (“1159”; 42.4±14.0% vs. 31.2±4.8%; n=15, p = 0.006; Figure 2A-B), alongside a higher proportion of glucagon-positive (GLC^+^) cells (19.4±6.7% vs. 13.6±4.2%; n=9, p = 0.04; Figure 2C).

**Figure 2.**
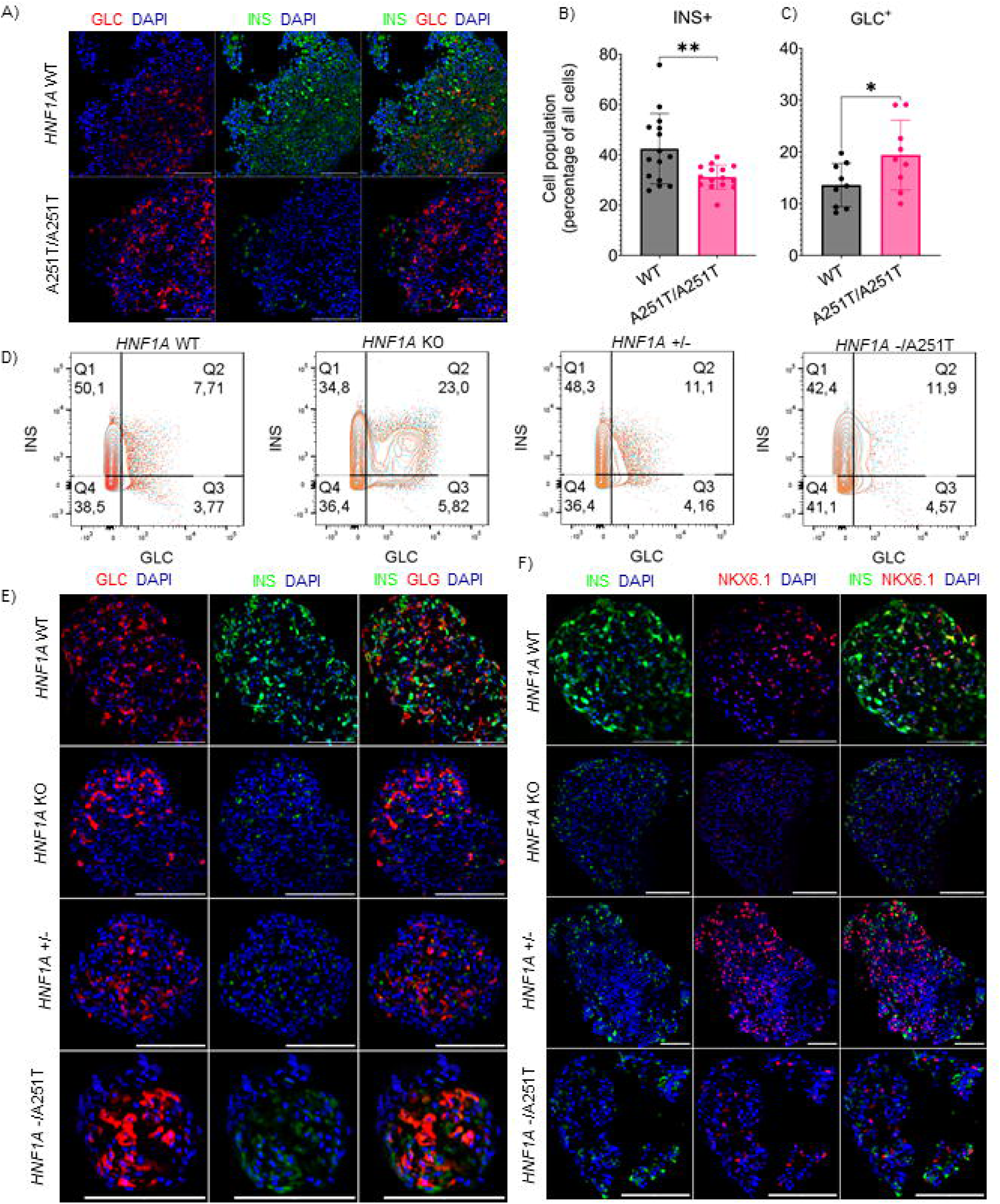
Altered β- and alpha-cell composition in homozygous *HNF1A* p.A251T iPSC- and hESC-derived islet-like clusters. (A) Representative immunofluorescence images of stage 6 iPSC-derived islet-like clusters from a control line (1159) and a proband carrying the homozygous *HNF1A* p.A251T variant, stained for insulin (green) and nuclei (DAPI, blue), showing reduced β-like and increased alpha-like cell representation in A251T clusters. (B–C) Quantification of insulin-positive (INS) and glucagon-positive (GLC) cells in iPSC-derived clusters from control and A251T lines (mean ± SD, n = 9–15). (D) Flow cytometry analysis (FlowJo) of hESC-derived islet-like clusters from three independent differentiations (>1 × 10 cells/condition), comparing WT, *HNF1A*+/–, *HNF1A*–/A251T, and *HNF1A*–/– genotypes. (E) Representative immunofluorescence images of hESC-derived clusters stained for insulin (green), glucagon (red), and nuclei (DAPI, blue). (F) Representative staining of insulin and NKX6.1 showing double-positive INS /NKX6.1 β-like cells. All images acquired on a STELLARIS confocal microscope using a 20× dry objective. Scale bar of 100 μm is applicable to all images. Significance *p ≤ 0.05.

To assess gene dosage effects, we compared CRISPR/Cas9-edited lines. Flow cytometry confirmed reduced proportions of INS /GFP beta-like cells in *HNF1A*^+/–^ (48.3%) and *HNF1A*^A251T/–^ (42.4%) compared to WT (50.1%; Figure 2D, 3H). Immunostaining revealed significantly fewer INS cells in both *HNF1A*^A251T/–^ (25.7±5.8%) and *HNF1A*^−/–^lines (23.1±3.9%) compared to WT (54.8±16.8%; all p<0.0001; Figure 2E, 3B), consistent with the reduction observed in patient-derived A251T iPSCs. A comparable increase in alpha-like cells proportion was observed in *HNF1A*^A251T/–^ (29.2±13.4%) and *HNF1A*^−/–^ clusters (29.3±8.6%), relative to WT (15.5±9.5%, Figure 2E, 3A). These findings demonstrate that the *HNF1A* p.A251T variant disrupts beta-cell differentiation and shifts the endocrine lineage balance toward alpha-cell formation.

Immunostaining revealed a higher proportion of INS /GLC double-positive cells in *HNF1A*^A251T/–^ clusters (14.8 ± 4.1%) compared to *HNF1A*^+/–^ controls (7.0 ± 4.7%; p = 0.006), similar to *HNF1A*^−/–^ (14.5 ± 5.1%) and elevated relative to WT (7.5 ± 4.7%) (Figure 2E, 3C). These double-positive cells indicated impaired beta-cell specification. NKX6.1 beta-like cells were significantly lowered only in *HNF1A*^−/–^clusters (13.2±7.0% vs. WT: 25.8 ± 8.8%; p=0.01), but remained stable in *HNF1A*^A251T/–^ (15.7 ± 5.8%) and *HNF1A*^+/–^ lines (19.8 ± 9.9%, Figure 2F, 3D-E), suggesting that partial *HNF1A* loss does not compromise *NKX6.1* expression. qPCR on sorted INS /GFP cells demonstrated elevated glucagon mRNA expression in *HNF1A*^A251T/–^ and *HNF1A*^−/–^ clusters (AUC WT: 1.0±0.1; 1.6±0.5; KO: 1.7±0.6; *HNF1A*^+/–^: 1.3±0.3; *HNF1A*^A251T/–^), indicating persistent alpha-cell transciptional programs (Figure 3F).

**Figure 3.**
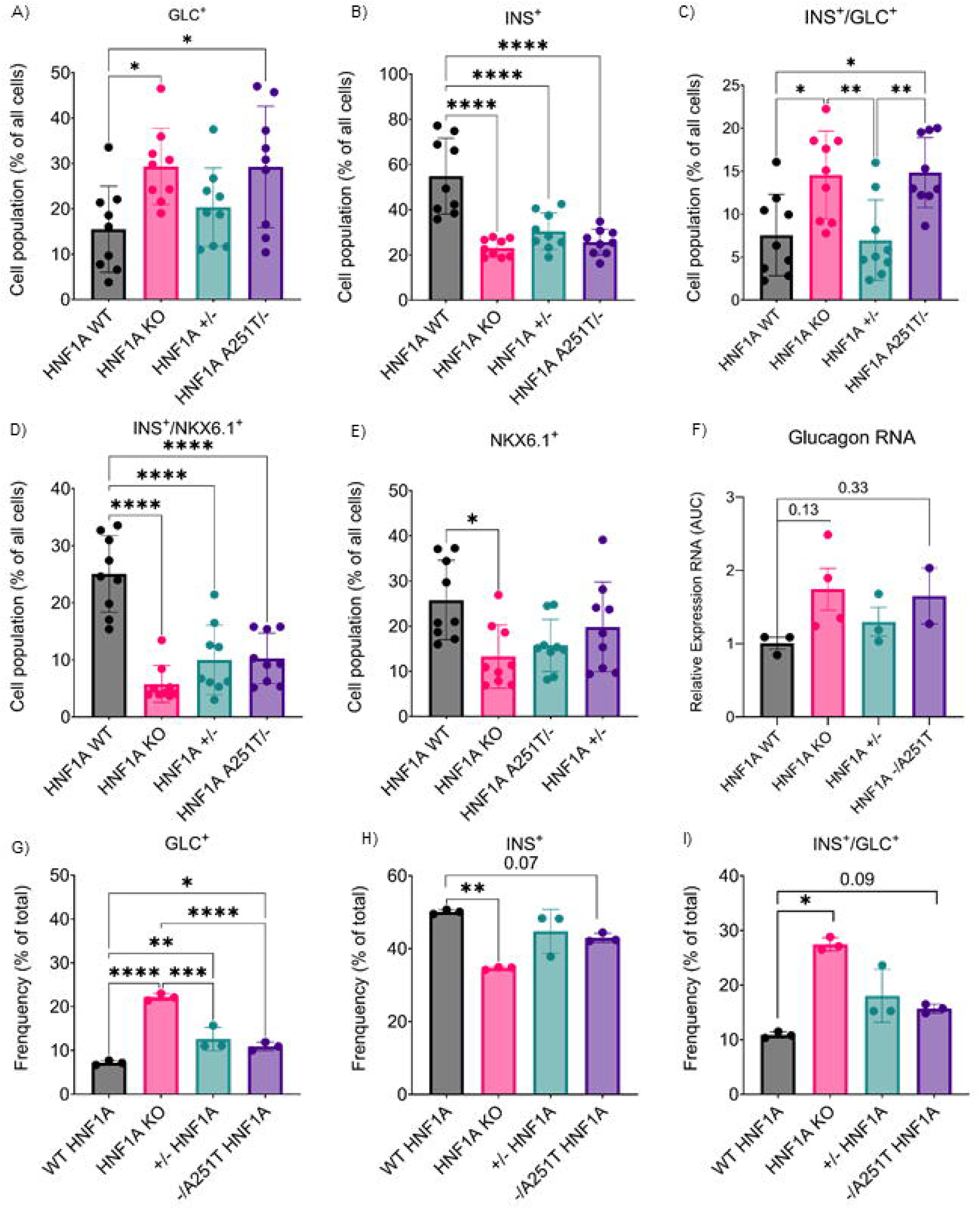
Quantitative analysis of altered β- and alpha-cell identity in hESC-derived islet-like *HNF1A* WT, *HNF1A*^+/–^, *HNF1A*^A251T/–^, and *HNF1A*^−/–^ clusters. (A–E) Quantification of endocrine subpopulations across WT, *HNF1A*^+/–^, *HNF1A*^A251T/–^, and*HNF1A*^−/–^ clusters, showing: (A) GLC alpha-like cells, (B) INS β-like cells, (C) GLC /INS double-positive cells, (D) INS /NKX6.1 double-positive β-like cells, and (E) NKX6.1 nuclei. Histograms represent mean ± SD from n = 9 biological replicates. (F) Quantitative RT-PCR for GLC mRNA in GFP-sorted INS β-like cells from each genotype, showing upregulation in *HNF1A*^A251T/–^ and *HNF1A*^−/–^ cells compared to controls (WT). (G–I) Flow cytometry–based quantification (FlowJo) of GLC, INS, and INS /GLC cell populations across genotypes. *p ≤ 0.05; non-significant trends are indicated with exact p-values.

### Impaired GSIS in HNF1A p.A251T beta-like cells is partially rescued by sulfonylureas

Dominant *HNF1A* mutations are known to impair GSIS in both human and murine models, including HNF1A KO hESC-derived beta-like cells^16,27^. To determine whether the recessive p.A251T variant causes a similar defect, insulin secretion was measured in iPSC-derived and CRISPR/Cas9-edited beta-like cells under low and high glucose, and with the glucagon-like peptide-1 (GLP-1R) receptor agonist, Exendin-4 or glibenclamide (GLB, sulphonylurea). Homozygous A251T iPSC-derived beta-like cells showed reduced responses to 16.7 mM glucose (0.05% vs. 0.20% of insulin content; p<0.001), Exendin-4 (10 nM, p < 0.0001), and GLB (10 μM, p=0.001) (Figure 4A). Similarly, CRISPR/Cas9-edited *HNF1A*^A251T/–^ and *HNF1A*^-/-^ hESC-derived clusters showed impaired GSIS (WT vs. KO, p = 0.02, WT vs *HNF1A*^A251T/–^ p=0.01; Figure 4B), with GLB partially restoring secretion (A251T: p=0.001; Fig.4C). These findings align with the proband’s clinical response to sulfonylurea^6^.

**Figure 4.**
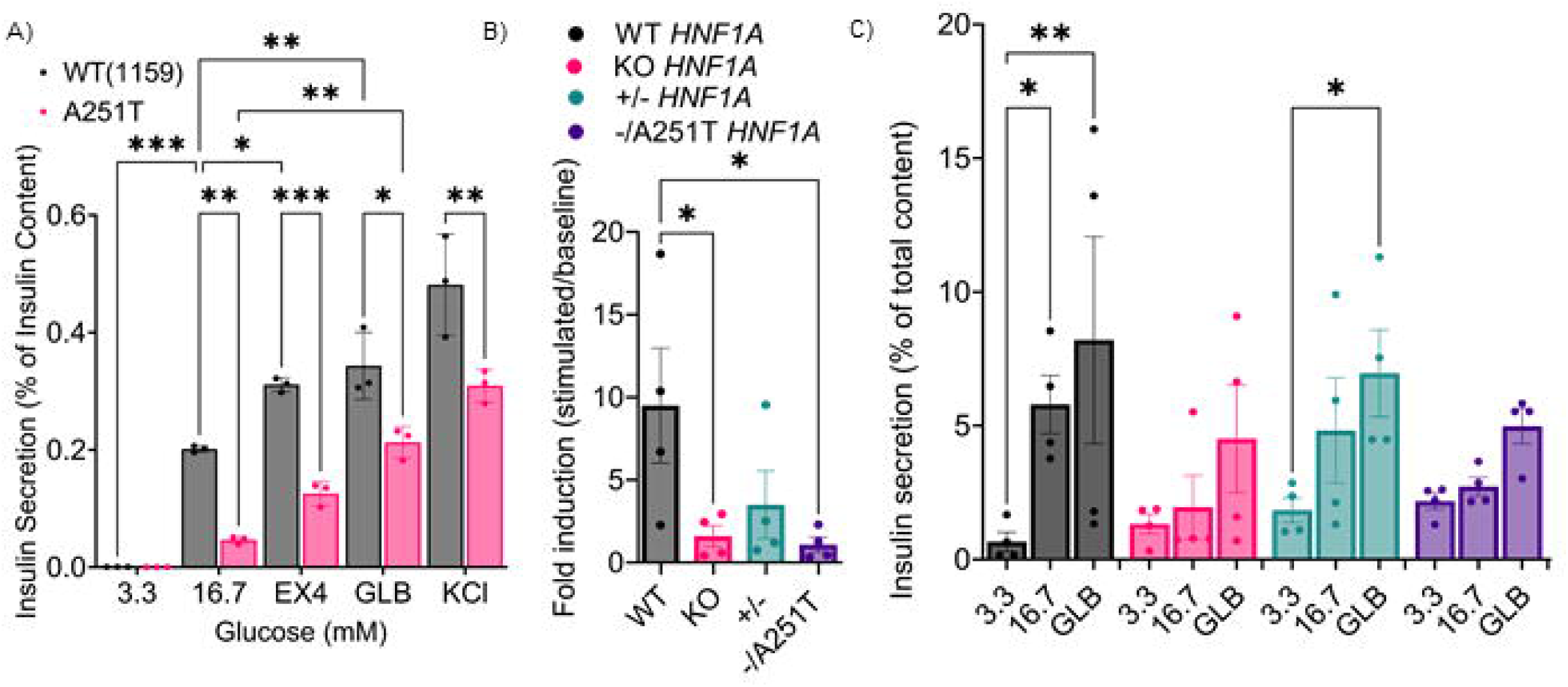
Impaired glucose-stimulated insulin secretion in *HNF1A* p.A251T β-like cells is partially rescued by glibenclamide. (A) Percentage of insulin secreted relative to total insulin content from iPSC-derived β-like clusters (WT and A251T/A251T) under basal glucose (3.3 mM), high glucose (16.7 mM), high glucose plus Exendin-4 (GLP-1 receptor agonist, 10 nM), high glucose plus glibenclamide (GLB, 10 μM), and low glucose plus KCl conditions. Insulin was measured using the Insulin Ultra-Sensitive HTFR kit. (B–C) Glucose-induced fold change in insulin secretion and percent secretion relative to total content in hESC-derived β-like clusters across genotypes: WT, *HNF1A*^+/–^, *HNF1A*–/A251T, and *HNF1A*^−/–.^ Data represent mean ± SD from three independent differentiations. Significance *p ≤ 0.05.

### Reduced Ca² responses and coordination in HNF1A p.A251T beta-like clusters

Ca² responses to 11 mM glucose were significantly lowered in *HNF1A*^A251T/–^ hESC-derived beta-like clusters compared to WT (p = 0.02; Figure 5A–D). The cumulative Ca² response (area-under-the-curve, AUC) in *HNF1A*^A251T/–^ clusters was comparable to *HNF1A*^−/–^, indicating impaired glucose-induced depolarization and Ca² influx (Figure 5D). However, responses in *HNF1A*^A251T/–^ and *HNF1A* ^+/–^ lines did not differ significantly (Figure 5D–E), suggesting that p.A251T alone does not exacerbate the calcium signaling defect beyond partial *HNF1A* loss. These results support a partial defect in glucose-induced Ca² signaling and insulin secretion, consistent with the variant’s recessive inheritance and mild phenotype.

**Figure 5.**
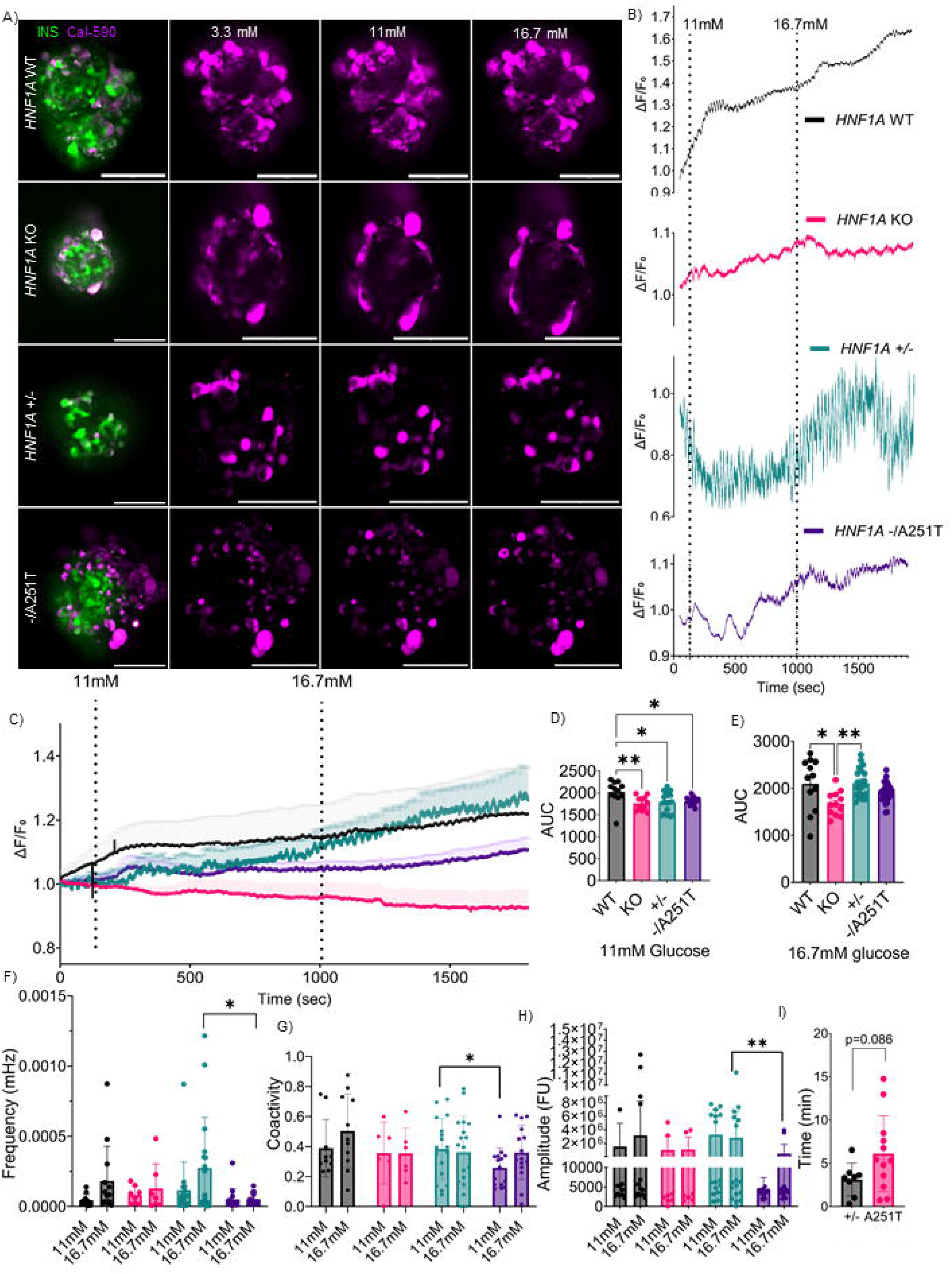
Impaired glucose-stimulated Ca²□ dynamics in *HNF1A* p.A251T β-like cells. (A) Cytosolic Ca^2+^ imaging of hESC-derived islet-like clusters pre-incubated in glucose- and glutamine-free DMEM containing Cal-590, imaged in real-time using a Zeiss Axiovert 2 spinning microscope; 20× dry objective. Baseline recordings were obtained at 3.3 mM glucose for 3 minutes, followed by sequential stimulation with 11 mM and 16.7 mM glucose for 15 minutes each. Scale bar of 100 μm is applicable to all. (B) Representative calcium fluorescence traces expressed as ΔF/F over time. (C) Mean Ca^2+^ response traces expressed as ΔF/F over time across genotypes (n=9). (D–E) Quantification of the area under the curve (AUC) for Ca² responses at 11 mM (D) and 16.7 mM glucose (E). (F) Oscillation frequency (mHz), (G) β-cell coactivity, (H) Signal amplitude (fluorescence units), and (I) Time to initial response (minutes), each measured at 11 mM and 16.7 mM glucose for the following genotypes: WT, *HNF1A* /, *HNF1A* /, and *HNF1A* /A251T. Data are shown as mean ± SD from at least three independent experiments. Statistical comparisons were performed using two-way ANOVA with Tukey’s post hoc test (for multiple groups), unpaired t-tests (for *HNF1A* / and *HNF1A* /A251T comparisons), or Kruskal–Wallis test where appropriate. Statistical significance was defined as p < 0.05. Movies corresponding to WT, *HNF1A* /, *HNF1A* /, and *HNF1A* /A251T are provided (Movie1 through Movie 4, respectively).

Compared to *HNF1A*^+/−^ cells, *HNF1A*^A251T/–^ cells failed to show an increased oscillation frequency in response to 16.7 mM glucose (p = 0.01; Fig. 5F). At 11 mM glucose, pairwise co-activity values were significantly lower in *HNF1A*^A251T/–^ clusters (mean=0.2) compared to *HNF1A*^+/−^(mean=0.3; p=0.03; Figure 5G), indicating impaired intercellular synchronicity. In addition, oscillation amplitude was consistently reduced in *HNF1A*^A251T/–^ clusters compared to *HNF1A*^+/−^ cells (p=0.004; Figure 5H). *HNF1A*^A251T/–^clusters also showed a trend towards delayed activation upon stimulation at 11 mM glucose (median=6.1±4.4 min.) compared with *HNF1A*^+/−^ cells (3.1± 0.19 min.; p = 0.09; Figure 5I), suggesting slower recruitment of responsive cells. Thus, *HNF1A* p.A251T impairs beta-cell network function by disrupting beta cell connectivity, likely contributing to defective insulin secretion^28^.

### Transcriptomic alterations in HNF1A p.A251T cells indicate immature beta-cell states and impaired secretion

To determine whether the *HNF1A* p.A251T variant induces transcriptomic changes comparable to complete *HNF1A* loss, we performed bulk RNA-seq on CRISPR/Cas9-edited and control cells followed by gene set enrichment analysis.

Interestingly, transcriptomic analysis revealed gene subsets differentially sensitive to HNF1A dosage. One group (including *ALDOB*, *HABP2*, *LIPC*, *AMKS4B*, *DPEP1*, *IL32*, *FABP1*, *LAMB3*, *SERPINA10*, and HSP family members) was specifically responsive to complete *HNF1A*^-/-^, but not in heterozygous or *HNF1A*^A251T/+^ lines (Figure 6A). This suggests that their expression requires full HNF1A activity, and may reflect functions linked to metabolic programs not maintained in beta-like cells under partial *HNF1A* loss.

**Figure 6.**
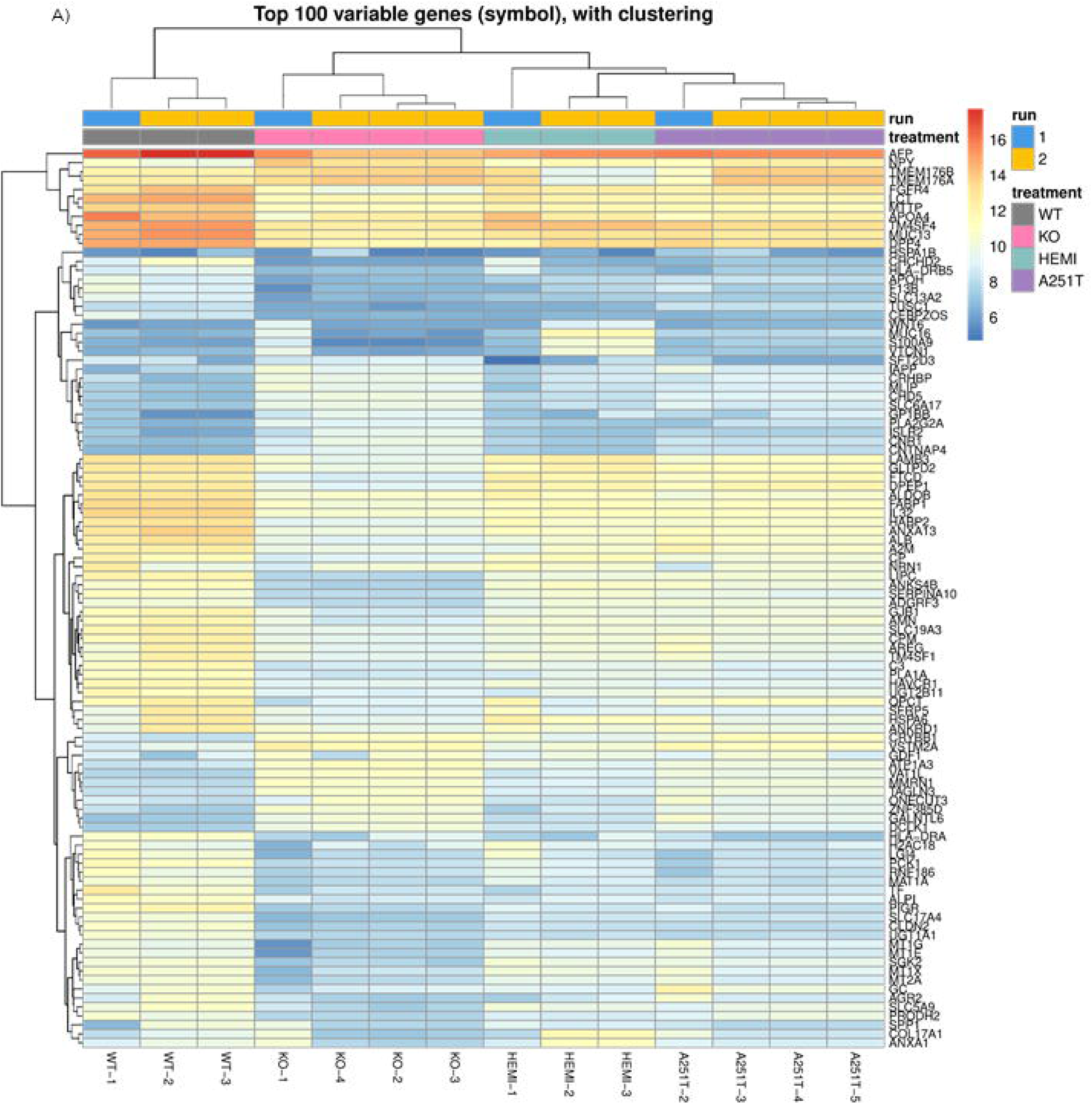
Transcriptomic clustering and heatmap of top variable genes across HNF1A genotypes. Heatmap displaying the top 100 most variable genes across all samples, based on variance-stabilized gene expression values and adjusted p-value filtering across ∼20,000 genes. Sample-wise hierarchical clustering and Euclidean distance metrics were applied to assess transcriptomic similarity among genotypes. Colors indicate genotype group: WT (gray), *HNF1A*^−/–^ (pink), *HNF1A*^+/–^ (turquoise), and *HNF1A* ^A251T/-^ (purple). Each group includes 3–4 biological replicates. Raw FASTQ reads were processed using the GenPipes v6.0.0 RNA-seq pipeline^21^ and transcript-level quantification was performed with kallisto v0.50.0 using the GRCh38 Ensembl v90 reference annotation^22^.

In contrast, a distinct set of genes, including *HSPA1B*, *CEBPZOS*, *HLA-DRA*, *PCK1*, *ANXA1*, *SLC5A9*, *SPP1*, and *MT1X/NT2A-* was reduced in *HNF1A*^+/–^ and *HNF1A*^A251T/–^cells, with no further decrease in KO cells. This indicates greater sensitivity to partial HNF1A insufficiency, consistent with haploinsufficiency (Figure 6A). Many of these genes are involved in stress response, cell signaling, and metabolic adaptation, suggesting they may be relevant to beta-cell maturation or resilience in stem cell-derived islet-like clusters.

Comparing KO vs WT beta-like cells, we observed broad pathway dysregulation, with 1,697 Gene Ontology (GO C5) pathways significantly altered (501 upregulated, 1,196 down-regulated) (Figure 7A, Supplementary Table 4). Down-regulated pathways were dominated by terms related to *lipid*, *fatty acid*, *steroid*, and *organic acid metabolism*, as well as *epithelial polarity/brush border organization* and *cytokine-mediated signaling*, consistent with loss of mature beta-cell metabolic and epithelial identity. In contrast, upregulated pathways were enriched for gene sets annotated as *synapse organization*, regulation of *trans-synaptic signaling*, *postsynaptic membrane/specialization*, and *axon development*, reflecting changes in ion-channel, Ca²-handling, and vesicle trafficking gene networks that support stimulus–secretion coupling (Figure 7A, Supplementary Table 4).

**Figure 7.**
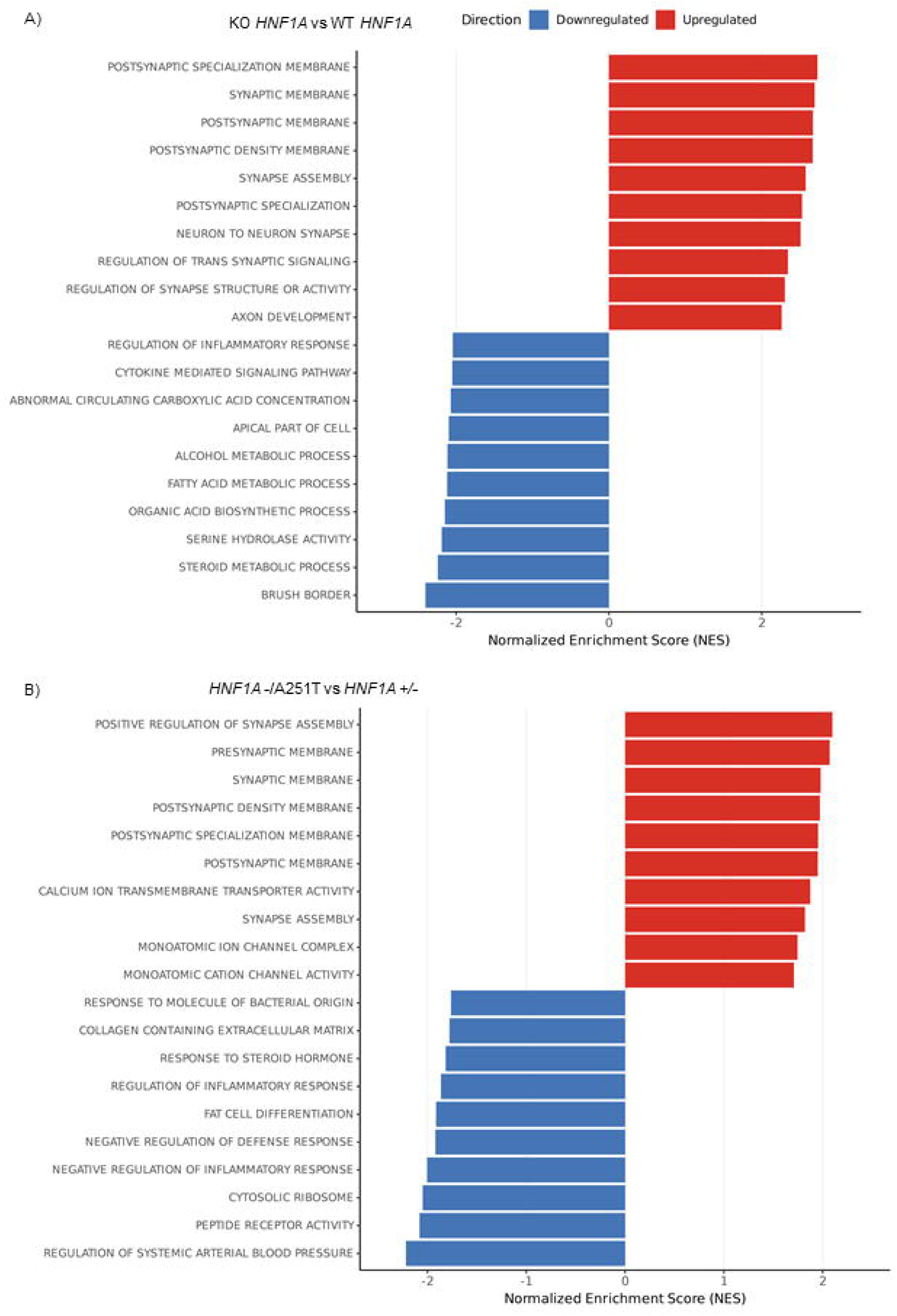
Gene et Enrichment Analysis reveals distinct transcriptional programs in HNF1A A251T/- β-like cells. (A) Top 10 significantly enriched Gene Ontology C5 biological process pathways identified by fast gene set enrichment analysis in *HNF1A* / versus WT β-like cells. Downregulated pathways in *HNF1A* / are shown in blue; upregulated pathways are shown in red. (B) Top 10 enriched GO C5 pathways in *HNF1A* ^A251T/-^ versus *HNF1A* / β-like cells. Blue indicates pathways downregulated in A251T relative to hemizygous; red indicates upregulated pathways in A251T. Gene set enrichment analysis was performed using the fgsea R package (v1.34.2) and over-representation analysis of gene ontology terms was conducted with gProfiler2 (v0.2.3)^25^.

In the *HNF1A*^A251T/–^ vs *HNF1A*^+/–^ comparison, the number of significantly enriched pathways was lower (420 GO C5: 99 up-regulated, 321 down-regulated) (Figure 7B, Supplementary Table 5), indicating a more restricted transcriptional impact than complete knockout, as may be predicted. Upregulated pathways again involved synapse-and ion-channel-related ontologies, including *monoatomic ion channel complex*, *calcium ion transmembrane transporter activity*, *synapse assembly*, and *postsynaptic membrane/specialization*. This pattern is consistent with a selective alteration of membrane excitability and Ca²-handling genes in *HNF1A*^A251T/–^ beta-like cells. Downregulated pathways included *peptide receptor activity*, *cytosolic ribosomal gene sets*, *collagen-containing extracellular matrix*, *response to steroid hormone*, and multiple terms related to regulation of inflammatory and defense responses, suggesting reduced biosynthetic capacity and altered interaction with extracellular and stress signals (Figure 7B, Supplementary Table 5).

Differential expression analysis revealed that compared with WT, *HNF1A* KO showed 7,177 DEGs (padj<0.05), whereas *HNF1A*^+/–^ exhibited 4,159 DEGs (Figure 8A). The *HNF1A*^A251T/–^genotype showed 6,834 DEGs compared to WT, closely matching the KO. Direct comparison between *HNF1A*^A251T/–^ and *HNF1A*^+/–^ identified 2,061 DEGs, indicating that the p.A251T variant induces transcriptional alterations beyond simple haploinsufficiency. Hierarchical clustering based on the top variable genes placed *HNF1A*^A251T/–^ samples transcriptionally between *HNF1A*^+/–^ and *HNF1A*^-/-^ but consistently distinct from *HNF1A*^+/–^(Figure 8A-B).

**Figure 8.**
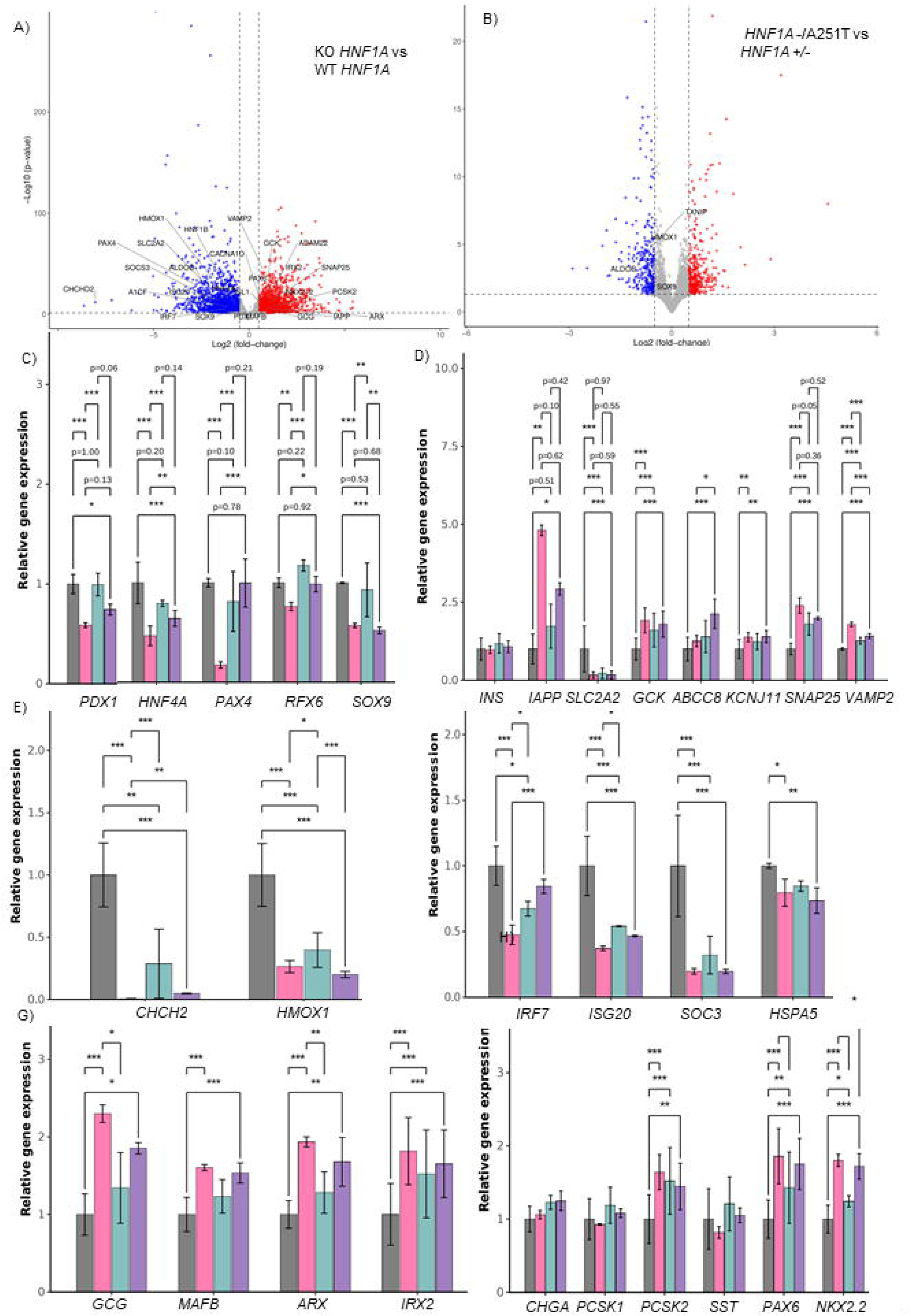
Transcriptomic profiling reveals genotype-specific changes in gene expression *HNF1A* A251T^/-^. (A–B) Volcano plots showing differential gene expression between (A) *HNF1A* WT) and *HNF1A*^-/-^ and (B) *HNF1A* ^A251T/-^ and *HNF1A*^+/–^ hESC-derived islet-like clusters. Gene expression counts were quantified using kallisto v0.50.0 against the GRCh38 Ensembl v90 transcriptome annotation. Differential expression analysis was performed using DESeq2 v1.48.1, adjusting for sequencing batch effects. (C) Expression levels of key β-cell identity and function genes. (D) Stem cell-to-beta-cell differentiation markers. (E) Mitochondrial and metabolic genes. (F) Stress and inflammation-associated transcripts. (G) Alpha-cell identity and function markers *GLC*. (H) Pan-endocrine and islet-enriched genes. Data represent normalized gene counts (or transformed counts if applicable); statistical significance thresholds were based on adjusted p-values< 0.05.

The *HNF1A* KO vs WT comparison revealed a strong shift away from mature β-cell gene programs toward a mixed neuroendocrine–immature islet state. The top upregulated genes were enriched for neuropeptides, ion channels, and synaptic/adhesion regulators (*NPY, GAP43, GRID2, SLITRK5, GABRA1/3, KCNA1/2, NTRK3*) (Figure 8A, Supplementary Table 6), while downregulated genes involved glucose metabolism, secretory function, and beta-cell identity and maturation (*ALDOB*, *PCK1*, *TXNIP*, *INSIG1*, *IGF2*, *CCND2*) (Figure 8A, Supplementary Table 7), consistent with beta-cell dedifferentiation accompanied by activation of alternative endocrine–neuronal transcriptional pathways.

The *HNF1A*^A251T/–^ vs *HNF1A*^+/–^ comparison showed a similar shift, though milder. Upregulated genes included neuronal/endocrine adhesion molecules and ion channels (*CNTNAP4, GRIA1, KCNA1/2, CNR1, PANX3, GAP43*) (Figure 8B, Supplementary Table 8), while the top down-regulated genes again involved beta-cell metabolic and stress-response functions (*PCK1, ALDOB/ALDOC, GADD45B, TXNIP, CEBPA, SMAD3*) (Supplementary Table 9).

Together, these signatures indicate that both complete *HNF1A* loss and the p.A251T variant weaken beta-cell identity and promote acquisition of a neuroendocrine-like transcriptional state, with A251T acting as a partial-loss allele.

### Distinct transcriptomic remodeling in HNF1A p.A251T beta-like cells reveals preserved exocytotic programs despite impaired glucose uptake and stress signaling

To further evaluate the functional consequences of *HNF1A* perturbation, we analyzed DEGs across beta-cell relevant categories, selected based on established HNF1A targets and published datasets of endocrine differentiation from hESC-derived models^19,29^. Transcriptomic profiling revealed that *HNF1A*^A251T/–^ and *HNF1A*^+/–^ beta-like clusters shared a broad transcriptional remodeling of *glucose sensing*, *exocytosis*, and *endocrine identity* pathways, partially overlapping with the *HNF1A*^−/–^ profile, yet with distinct features in the *HNF1A*^A251T/–^ line.

Notably, the glucose transporter *SLC2A2/GLUT2* gene was significantly downregulated across all HNF1A-deficient lines, suggesting a shared impairment in glucose uptake (log FC −2.32, −2.09, −2.12, respectively; all padj <2×10 ¹; Figure 8D). In contrast, genes involved in vesicle docking and stimulus–secretion coupling, including *ABCC8, KCNJ11, STX1A, SNAP25, VAMP2* and *EXOC7*, were significantly upregulated in *HNF1A*^A251T/–^ cells (e.g. *ABCC8* log FC 1.03, padj 6.5×10; *SNAP25* log FC 0.97, padj 1.3×10; Figure 8D), and more so than in the *HNF1A*^+/–^.

Several canonical beta-cell transcription factors were significantly down-regulated. *HNF4A, PDX1,* and *HNF1B* were decreased in *HNF1A*^A251T/–^ compared to WT (log FC −0.51, −0.38, −0.69; padj 2.9×10, 0.023, 4.2×10; Figure 8C-D), with even greater suppression observed in *HNF1A*^−/–^line (e.g., *HNF4A* log FC −1.03, padj 2.4×10 ¹³). In addition, mitochondrial and oxidative stress regulators were disrupted. *HMOX1* and *CHCHD2* were downregulated across all HNF1A-compromised genotypes, with maximal repression of *CHCHD2* in *HNF1A*^−/–^cells (log FC −8.05, padj 1.1×10 ¹²; Figure 8E). Interestingly, *CHCHD2* expression was partially restored in *HNF1A*^A251T/–^ compared to KO (log FC 0.05, padj 4.6×10 ³) and substantially increased in *HNF1A*^+/–^ vs KO (log FC 3.52, padj 8.3×10), whereas *HMOX1* remained more suppressed in *HNF1A*^A251T/–^ than in the *HNF1A*^+/–^ line (log FC −0.72, padj 2.9×10, Figure 8E).

### HNF1A p.A251T promotes an alpha-cell–biased endocrine lineage

Markers of alpha-cell identity were consistently up-regulated across *HNF1A*-deficient lines, indicating a progressive shift away from a beta-cell fate. The alpha determinant *ARX* was significantly increased in *HNF1A*^A251T/–^ cells (log FC 0.58, padj=0.002, Figure 8G) and *HNF1A*^−/–^ cells (log FC 0.91, padj=1.4×10) compared to WT, but reduced in *HNF1A*^+/–^ cells relative to KO (log FC −0.53, padj=0.006, Figure 8G). A similar trend was observed for glucagon which was elevated in *HNF1A*^A251T/–^ cells (log FC 0.76, padj=0.01) and KO (log FC 1.16, padj = 2.5×10), but decreased in *HNF1A*^+/–^vs KO (log FC −0.65, padj=0.03, Figure 8G). Additional alpha-cell-associated transcription factors, including *IRX2* and *MAFB*, were also up-regulated across all mutant lines compared to WT (*IRX2* log FC 0.63–0.85, padj=10 ¹³; *MAFB* log FC 0.57–0.68, padj=4.0×10; Figure 8G).

## Discussion

The present study provides new insights into the cellular mechanisms behind the pathogenicity of the recessive *HNF1A* p.A251T variant and its impact on beta-cell development and function, clarifying its role in this recessive form of monogenic diabetes. Using a multi-modal, cell-based approach, including functional assays in immortalized lines, patient-derived iPSCs homozygous for p.A251T, and CRISPR/Cas9-edited hESCs differentiated into pancreatic beta-like cells, we demonstrate that the p.A251T variant impairs beta-cell glucose responsiveness and disrupts endocrine lineage specification. Importantly, we describe a pipeline including stem cell-derived cells and in-depth functional and phenotypic analysis. This approach, deployed for the first time here to explore a recessive variant, has allowed us to identify subtle, but functionally significant, effects. Demonstrating the power of this approach for precision medicine, we show preserved responses to sulphonylureas, as well as a to GLP-1R agonists in patient-derived cells.

Compared to our earlier report^6^, DNA binding assays in cell lines, using an alternative protocol (Methods), revealed a more modest (non-significant) effect for *HNF1A* p.A251T. Nevertheless, we recapitulated our findings on unaltered transactivation capacity of the A251T mutant protein (Figure 1). This is in marked contrast to dominant-negative mutations such as *HNF1A* p.P112L or p.T260M^9^. Interestingly, *HNF1A* p.A251T demonstrated significantly reduced nuclear localization *versus* the WT protein, likely contributing to its functional impairment. Again, the change was more modest than described^7,30^ for dominant mutations (eg. *HNF1A* p.P112L, p.T260M^9^).

Given the limitations of *in vitro* studies^31–34^, which usually involve overexpressed proteins in non-beta cell systems and preclude studies of islet cell development, we generated iPSCs from a proband homozygous for *HNF1A* p.A251T and CRISPR-edited hESCs differentiated into pancreatic beta-like cells.

*HNF1A*^+/–^ hESC-derived and patient-specific iPSC-derived beta-like cells carrying the homozygous p.A251T variant exhibited impaired GSIS, which was partially rescued by glibenclamide, reflecting the proband’s clinical response^6^. Isogenic CRISPR/Cas9 models confirmed this as a recessive defect: beta-like cells with *HNF1A*^A251T/–^ or *HNF1A*^−/–^genotypes were glucose-unresponsive, while *HNF1A*^+/–^ lines showed preserved insulin secretion. This convergence between clinical and *in vitro* responses underscores the utility of patient-specific and genome-edited stem cell models in variant classification.

In addition to impaired insulin secretion, *HNF1A*^A251T/–^ beta-like clusters exhibited reduced glucose-stimulated Ca^2+^ oscillation amplitude, frequency, and co-activity, resulting in attenuated network connectivity at 11 mM glucose. However, these defects were milder than those observed in *HNF1A*^−/–^ cells, indicating that the p.A251T variant confers a hypomorphic phenotype, with partial preservation of function.

Transcriptomic profiling further clarified the molecular impact of the *HNF1A* p.A251T variant. Core beta-cell transcription factors, including *NKX6.1*, remained largely preserved in *HNF1A*^A251T/–^ beta-like cells, yet key functional gene sets were perturbed. Genes involved in glucose transport (*SLC2A2/GLUT2*), insulin exocytosis (*STX1A, VAMP2*), ion channel activity (*ABCC8, CACNA1D, KCNJ11*), and oxidative stress response (*HMOX1, TXNIP*) were differentially expressed. The coordinated reduction in glucose-stimulated insulin secretion and calcium signaling in *HNF1A*-deficient beta-like cells is likely driven by impaired glucose sensing and stimulus-secretion coupling. For example, upregulation of *ABCC8*, *SNAP25* and *KCNJ11* may reflect compensatory responses to impaired depolarization and contribute to altered first-phase insulin release and oscillation frequency, respectively, while *GLUT2* downregulation could limit glucose responsiveness at physiological concentrations^35–37^. Notably, these transcriptional changes were derived from bulk RNA-seq of mixed endocrine clusters, and shifts in cell-type proportions-particularly increased alpha-like cells - may contribute to the observed expression patterns.

*HNF1A* p.A251T acts as a hypomorphic allele with recessive transmission in regards to MODY: mono-allelic inheritance is associated with later-onset diabetes^6,38^. Thus, quantitative lowering of HNF1A activity elicits beta-cell dysfunction by selectively impairing glucose sensing, stimulus-secretion coupling, and endocrine specification, without abolishing beta-cell identity. These results support the classification of p.A251T as a pathogenic variant causing autosomal recessive HNF1A-MODY, with implications for diagnostic reclassification and precision therapy.

Interestingly, and extending earlier results^16,27^, we also show that HNF1A has a critical role in controlling intracellular Ca^2+^ dynamics and, importantly, in the communication between individual cells *HNF1A*^A251T/–^ (“connectivity”).

### Limitations of the study

Since we were unable to generate cells homozygous for *HNF1A* p.A251T, as observed in the patients under study, we have relied here on comparing cells which were hemizygous for a null mutation in *HNF1A* with those bearing the same mutation plus the A251T variant. Whilst this approach is expected to provide a reasonable assessment of the impact of the loss of both alleles, future studies, perhaps based on newer gene editing technologies, will be required to confirm this assumption. We note also that future studies will be needed to explore lineage-restricted changes in gene expression profiles.

### Conclusions

This study provides new insights into the pathogenic mechanisms of the *HNF1A* p.A251T variant in the context of monogenic diabetes. More broadly, we reveal altered beta-cell connectivity as a mechanism through which perturbations in *HNF1A* impact islet function. Overall, our findings broaden understanding of HNF1A-related diabetes, emphasizing the importance of autosomal recessive *HNF1A* variants.

## Acknowledgments

I.C. was supported by a Diabetes UK studentship (BDA 18/0005934) to S.M. and G.A.R. G.A.R. was supported by a Wellcome Trust Investigator (w212625/Z/18/Z) Award, and MRC Programme grant (MR/R022259/1), Diabetes UK (BDA 16/0005485), an NIH-NIDDK project grant (R01DK135268) a CIHR-JDRF Team grant (CIHR-IRSC TDP-186358 and JDRF 4-SRA-2023-1182-S-N), CRCHUM start-up funds, and Innovation Canada John R. Evans Leader Awards (CFI 42649, CFI 46539). The authors thank the Facility for Imaging by Light Microscopy (FILM) at Imperial College London, which is partially supported by the Wellcome Trust (grant 104931/Z/14/Z) and the BBSRC (grant BB/L015129/1). MG is supported by a Postdoc Mobility Fellowship for the Swiss National Science Foundation (#225305). D.E. was supported by the Helmsley Charitable Trust. GO is the recipient of a postdoctoral FRQS scholarship (https://doi.org/10.69777/333390, https://doi.org/10.69777/376237). S.M. is funded by a Wellcome Trust Career Development Award (223024/Z/21/Z) and is supported by the NIHR Imperial Biomedical Research Centre. H.G.L. was supported by MRC core funding (MC-A652-5QA10) and NIHR Imperial BRC and is now supported by the NIHR GOSH BRC. The views expressed are those of the authors and not necessarily those of the NHS, the NIHR, or the Department of Health. Flow cytometry analyses were performed using the Cytometry Platform of the CRCHUM (Centre de recherche du Centre hospitalier de l’Université de Montréal), and we gratefully acknowledge PhD. Gaël Dulude, and MSc. Philippe St-Onge, as well as Dr Aurelie Cleret-Buhot (Plateforme d’imagerie cellulaire, CRCHUM) for her expert technical assistance. RNA-sequencing data analysis was performed by the Centre for the Integration and Analysis of Medical Data (CITADEL, CRCHUM, Montréal, Canada), and we gratefully acknowledge PhD. Pedro Bordignon, PhD. Maxime Caron, and the entire CITADEL team for their support and expertise. We thank Professor Alejandra Tomas (Imperial College London) for the provision of INS1(832/13) cells.

## Data availability statement

Bulk RNAseq data will be deposited at the GEO Database.

## Guarantor

GAR serves as the guarantor of this study.

## Author Contributions

I. Cherkaoui performed all experiments, with the exception of RNA-sequencing (RNA extraction was performed by I. Cherkaoui). Calcium imaging experiments were conducted jointly with G. Ostinelli.

M. Gasser contributed to stem cell culture and differentiation, including GSIS assays and qPCR. M. Gasser also assisted in total calcium analysis and AUC quantification.

Q. Du and D.M. Egli provided in-person training in β-cell differentiation at Columbia University’s Stem Cell Initiative as well as donation of the control iPSC line (1159) and HNF1A knockout hESC line.

C. Dion and H.G. Leitch provided training in iPSC reprogramming from skin biopsy samples.

G. Ostinelli contributed to calcium imaging acquisition and performed connectivity analyses.

D. Sachedina performed the 4 mm skin punch biopsy on the probands of the DIPS clinical study.

S. Misra co-supervised the project and led two clinical studies supporting this work: the *Studying Rare Diabetes Using Induced Pluripotent Stem Cells (DIPS)* study (IRAS ID: 280324; sponsor: Imperial College London), which enabled iPSC reprogramming from patient samples; and the *MODY in Young-Onset Diabetes in Different Ethnicities (MYDIABETES)* study (ClinicalTrials.gov ID: NCT02082132; sponsor: Imperial College London), through which the A251T proband was identified and recruited.

G.A. Rutter co-designed the study with S. Misra, co-supervised work, co-wrote the manuscript and provided funding.

