## Supplementary figures for "A recessive *HNF1A* p.A251T variant causes monogenic diabetes by altering islet cell development, insulin secretion and intercellular connectivity"

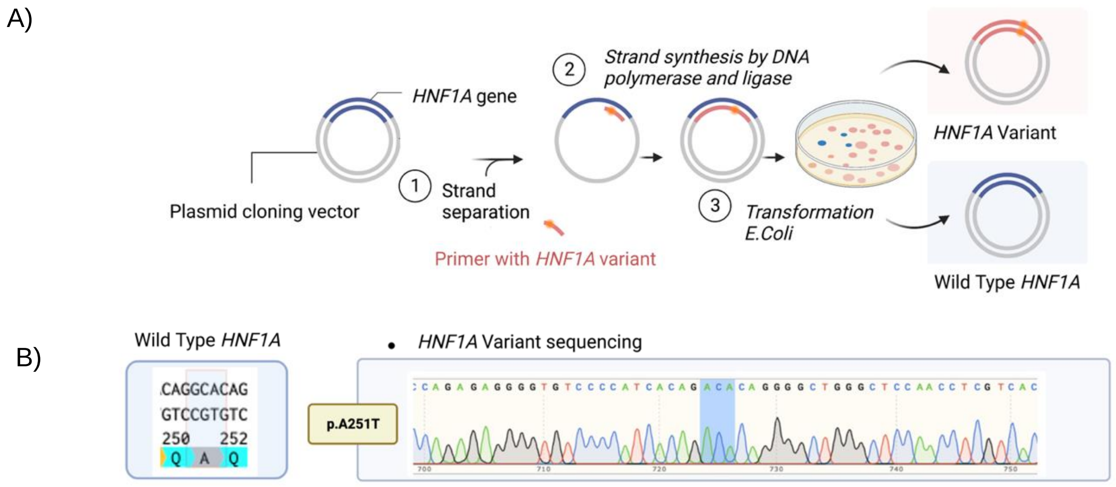
A)
HNF1A gene
Plasmid cloning vector
1 Strand separation
Primer with HNF1A variant
2 Strand synthesis by DNA polymerase and ligase
3 Transformation E.Coli
HNF1A Variant
Wild Type HNF1A
B)
Wild Type HNF1A
CAGGCACAG
GTCCGTGTC
250 252
Q A Q
p.A251T
HNF1A Variant sequencing
700
710
720
730
740
750

Fig S1.

Supplementary Figure 1. **Site-directed mutagenesis and plasmid validation**.

(A) Schematic representation of site-directed mutagenesis (SDM) workflow: PCR amplification using mismatch primers, enzymatic digestion with kinase/ligase/DpnI, and transformation into competent E. coli. (B) Representative sequencing results confirming successful introduction of *HNF1A* variant p.A251T compared with WT.

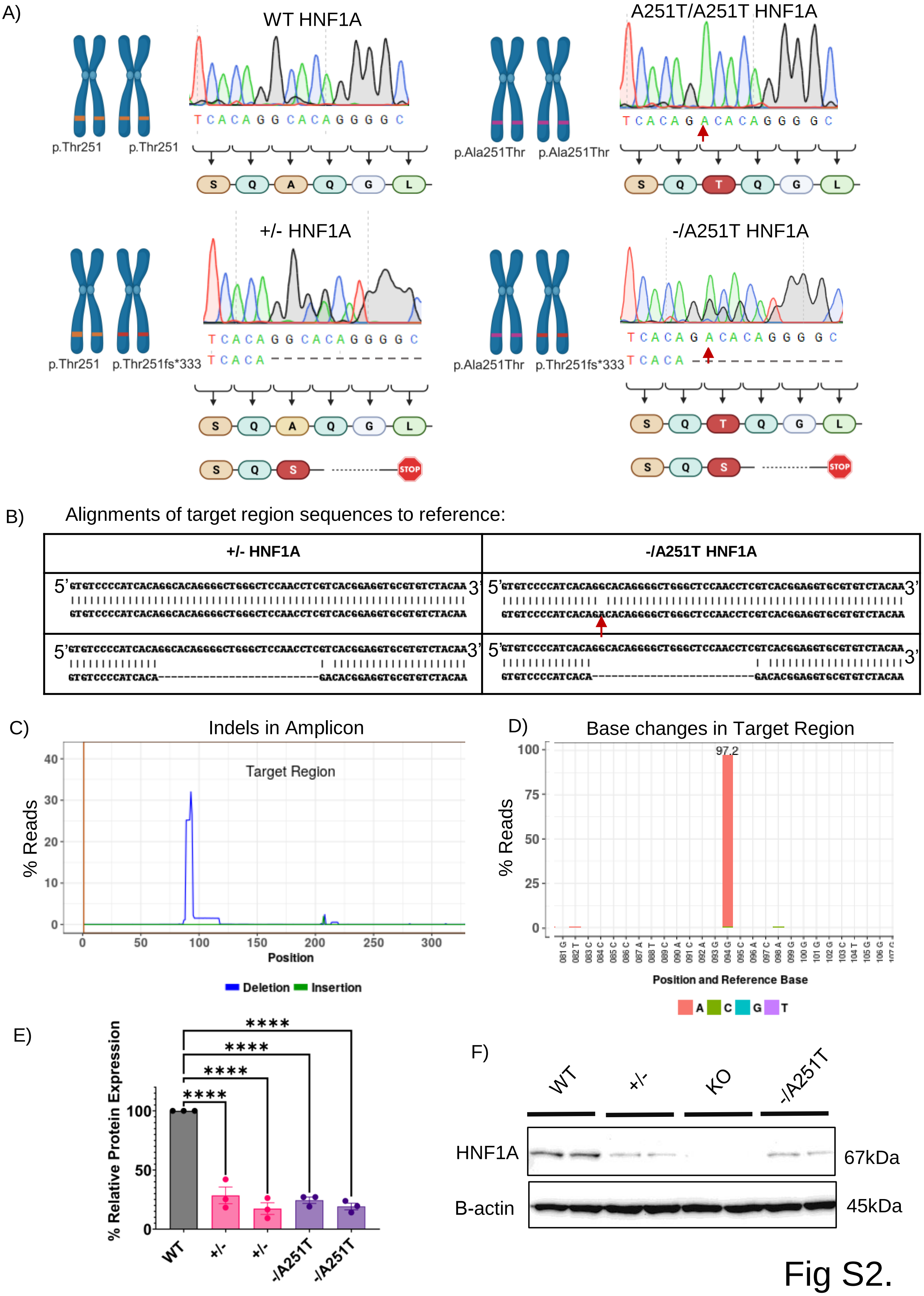
A)
WT HNF1A
p.Thr251
p.Thr251
A251T/A251T HNF1A
p.Ala251Thr
p.Ala251Thr
+/- HNF1A
p.Thr251
p.Thr251fs*333
-/A251T HNF1A
p.Ala251Thr
p.Thr251fs*333
STOP
B)
Alignments of target region sequences to reference:
+/- HNF1A
-/A251T HNF1A
C)
Indels in Amplicon
Target Region
% Reads
Position
Deletion
Insertion
D)
Base changes in Target Region
97.2
% Reads
Position and Reference Base
A
C
G
T
E)
% Relative Protein Expression
****
WT
+/-
-/A251T
F)
WT
+/-
KO
-/A251T
HNF1A
67kDa
B-actin
45kDa

Fig S2.

Supplementary Figure 2. **Generation and validation of HNF1A-edited stem cell lines**.

(A) Sanger sequencing of *HNF1A* alleles in WT, *HNF1A* p.A251T/p.A251T iPSCs, heterozygous *HNF1A* +/– (p.Thr251fs333) cells, and hemizygous *HNF1A* –/A251T lines. Chromatograms are shown alongside schematic representations of each allele and in silico predicted amino acid changes. (B) Next-generation sequencing (NGS) amplicon analysis confirming on- and off-target editing results (Genewiz). Aligned reads from *HNF1A* WT and +/– show a 25-bp deletion in one allele (p.Thr251fs333); comparison of WT and –/A251T reveals the same 25-bp deletion in one allele and the A251T single-nucleotide substitution in the other. (C) Quantification of edited reads showing the percentage of NGS reads carrying the 25-bp deletion at the sgRNA-targeted locus in the *HNF1A* +/– genotype. (D) NGS validation showing 97.2% base-pair concordance in the targeted HNF1A allele confirming successful introduction of the A251T SNP in cloned –/A251T cells (representative clone shown). (E) Quantification of HNF1A protein expression by Western blot, analyzed with Image Lab software and normalized to nuclear β-actin. Values are expressed relative to WT (100%). Two independent clones per genotype (+/–, –/A251T) were analyzed. (F) Representative Western blot showing HNF1A (67 kDa) expression in WT, +/–, –/A251T, and KO stem cell lines, with β-actin (~45 kDa) as a loading control.

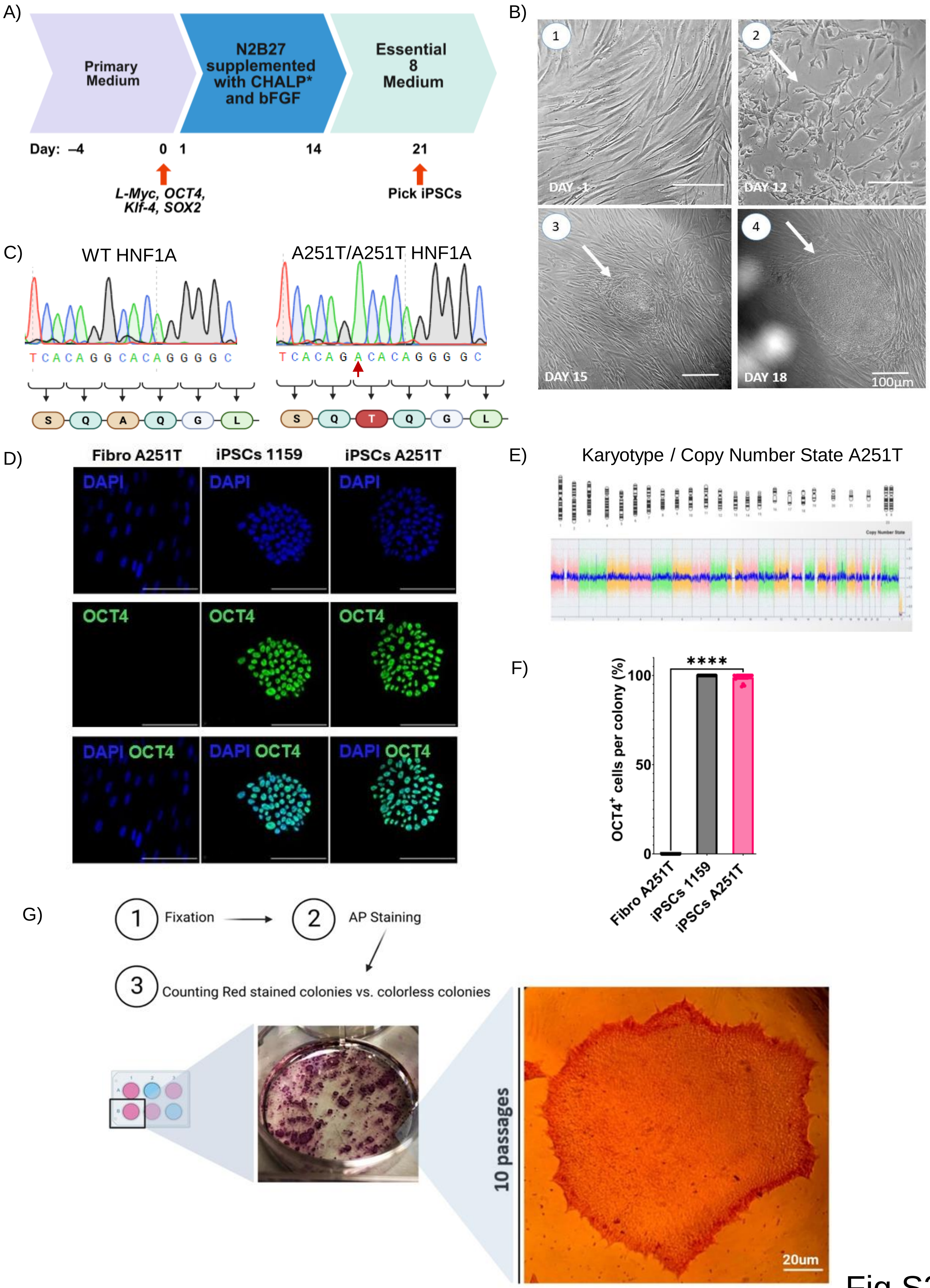
A)
Primary Medium
N2B27 supplemented with CHALP* and bFGF
Essential 8 Medium
Day: –4
0
1
14
21
L-Myc, OCT4, Klf-4, SOX2
Pick iPSCs
B)
1
2
3
4
DAY -1
DAY 12
DAY 15
DAY 18
100µm
C)
WT HNF1A
A251T/A251T HNF1A
T C A C A G G C A C A G G G G C
T C A C A G A C A C A G G G G C
S Q A Q G L
S Q T Q G L
D)
Fibro A251T
iPSCs 1159
iPSCs A251T
DAPI
OCT4
DAPI OCT4
E)
Karyotype / Copy Number State A251T
F)
OCT4+ cells per colony (%)
100
50
0
****
Fibro A251T
iPSCs 1159
iPSCs A251T
G)
1 Fixation
2 AP Staining
3 Counting Red stained colonies vs. colorless colonies
10 passages
20um

Fig S3.

Supplementary Figure 3. **Generation and characterization of *HNF1A* p.A251T patient-derived iPSCs**.

(A) Schematic representation of the episomal reprogramming protocol using OCT4, SOX2, KLF4, LIN28, and L-MYC. Fibroblasts were electroporated and cultured sequentially in N2B27 medium supplemented with CHALP factors, followed by Essential 8 medium until colonies appeared around day 21. (B) Phase-contrast images showing morphological changes during reprogramming from fibroblasts to iPSCs at days 1, 12, 15, and 18. White arrows indicate emerging iPSC colonies with typical pluripotent morphology. Scale bars: 100 µm. (C) Sanger sequencing chromatograms confirming homozygosity for the *HNF1A* p.A251T variant in patient-derived iPSCs and WT genotype in control iPSC line 1159. (D) Immunofluorescence staining of OCT-4 (green) and DAPI (blue) in fibroblasts, control iPSCs (1159), and A251T iPSCs. Nuclear OCT-4 localization is observed only in iPSCs, confirming pluripotency (Leica STELLARIS 8 confocal, ×40, oil immersion). Scale bar: 100 µm. (E) Karyotype analysis showing a normal 46, XX complement without detectable chromosomal aberrations. (F) Quantification of OCT-4–positive nuclei (% of total cells), analyzed using a custom macro (Imperial College Imaging Facility). Mann–Whitney U test, $p \leq 0.0001$ ($n = 15$). (G) Alkaline phosphatase (AP) staining of iPSC colonies at passage 10 showing uniform red staining in all clones, indicating robust AP activity and successful reprogramming.

A) Day of differentiation

Stage of differentiation

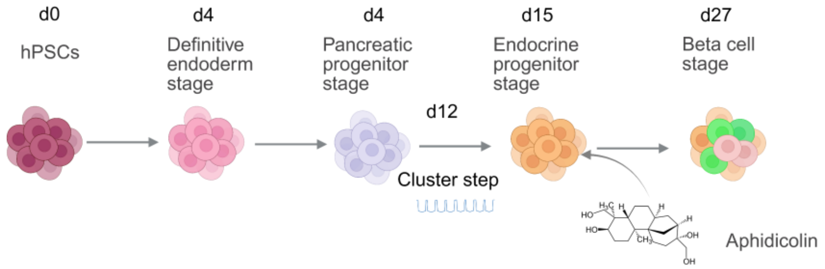


B)

| | | | | | | | |
|---|---|---|---|---|---|---|---|
| Small molecules | Ri | DE Kit Supp | KGF | RA LDN Cyclo KGF | EGF KGF | T3 RepSox KGF ZS UFH Ri XX APH | APH Ri |
| Basal medium | StemFlex | DE Kit | RPMI+ B27 | DMEM + GlutaMAX + B27 | | RPMI+B27 | RPMI+FBS+B27 |

C)

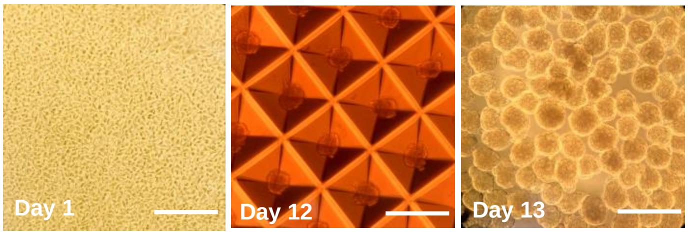


Fig S4.

Supplementary Figure 4. **Directed differentiation of human pluripotent stem cells into β-like cells**.

(A) Six-stage schematic of directed β-cell differentiation: (1) definitive endoderm, (2) primitive gut tube, (3) posterior foregut, (4) pancreatic progenitors, (5) endocrine progenitors, and (6) β-like cells. (B) Overview of signaling pathways and key small molecules used at each stage (e.g., Activin A, KGF, retinoic acid, LDN-212854, aphidicolin, and EGF). (C) Morphological progression from adherent hPSCs (day 0) to clustered endocrine progenitors (day 12) and mature β-like clusters (day 13 onward). Scale bars: 200 μm.

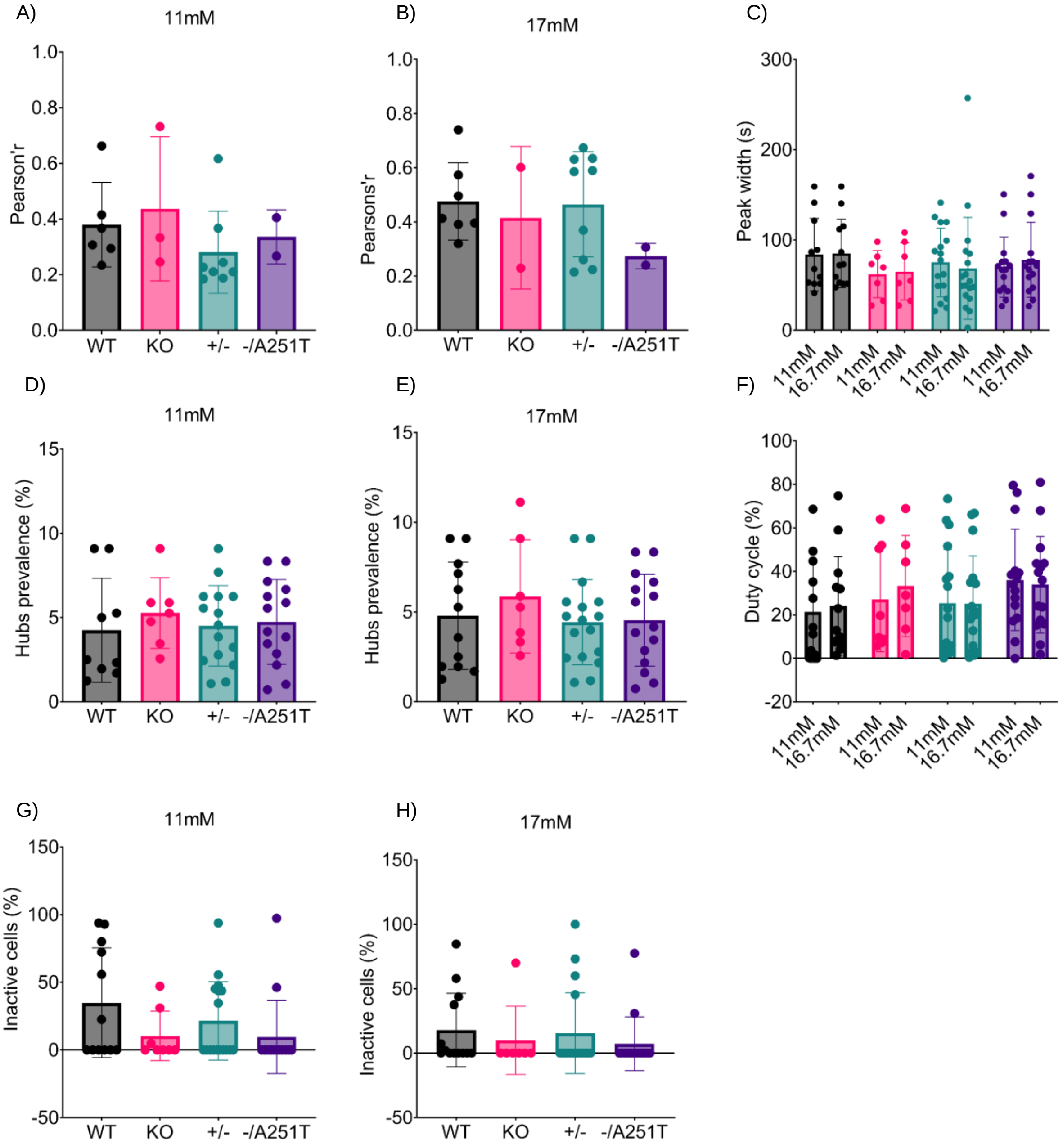
A)
11mM
Pearson'r
WT
KO
+/-
-/A251T
B)
17mM
Pearsons'r
C)
Peak width (s)
11mM
16.7mM
D)
Hubs prevalence (%)
E)
F)
Duty cycle (%)
G)
Inactive cells (%)
H)

Fig S5.

Supplementary Figure 5. **Analysis of $Ca^{2+}$ dynamics and connectivity**.

A-H analysed parameters as indicated. See methods for further details.

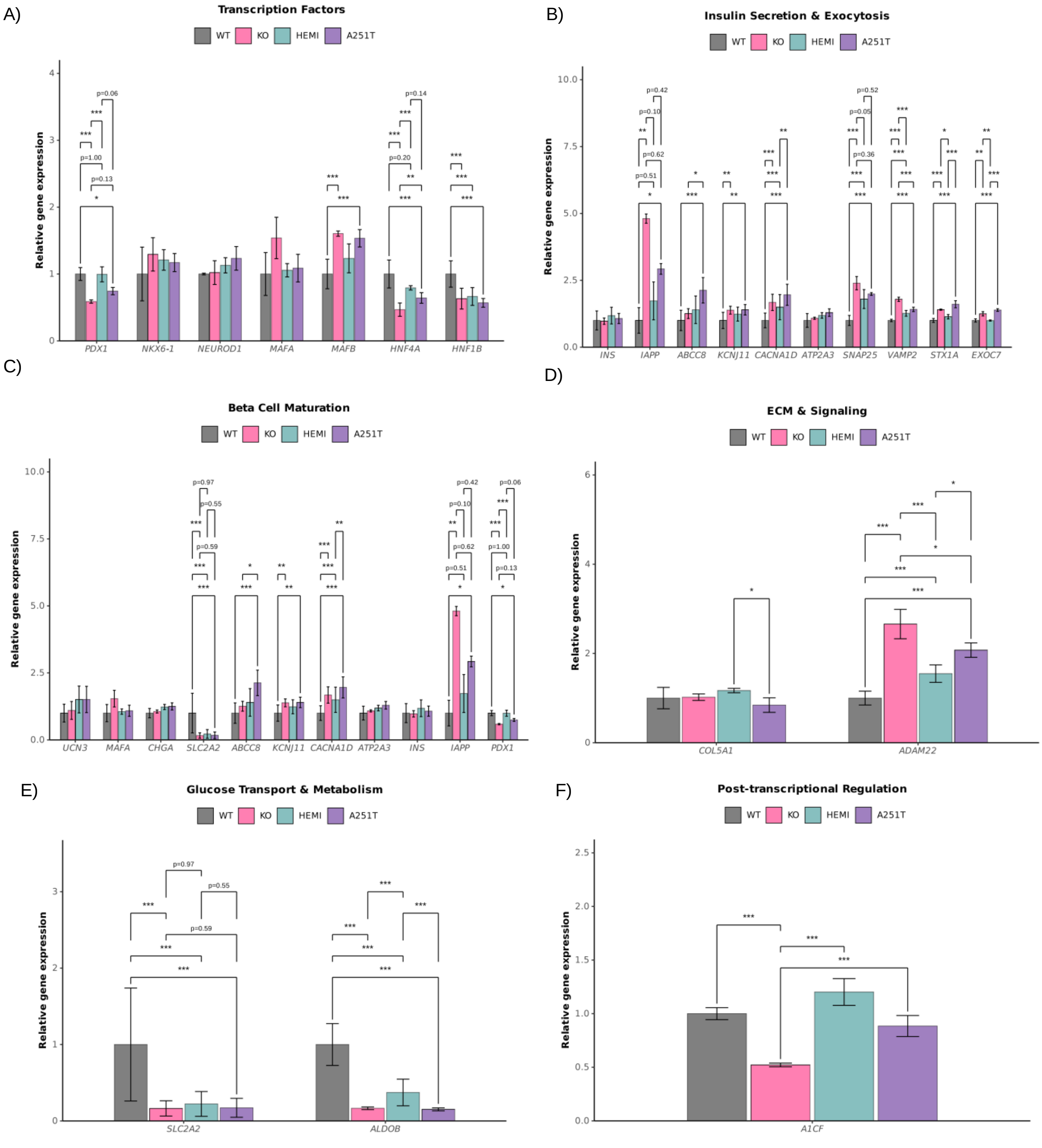
A)
Transcription Factors
WT
KO
HEMI
A251T
Relative gene expression
PDX1
NKX6-1
NEUROD1
MAFA
MAFB
HNF4A
HNF1B
B)
Insulin Secretion & Exocytosis
INS
IAPP
ABCC8
KCNJ11
CACNA1D
ATP2A3
SNAP25
VAMP2
STX1A
EXOC7
C)
Beta Cell Maturation
UCN3
MAFA
CHGA
SLC2A2
ABCC8
KCNJ11
CACNA1D
ATP2A3
INS
IAPP
PDX1
D)
ECM & Signaling
COL5A1
ADAM22
E)
Glucose Transport & Metabolism
SLC2A2
ALDOB
F)
Post-transcriptional Regulation
A1CF

Fig S6.

Supplementary Figure 6. **Histograms of gene expression changes across key molecular and biological function categories in stem cell–derived islet-like clusters.**

Histograms display $\log_2$ fold changes and adjusted p-values for representative genes within defined functional categories at Stage 6 of stem cell differentiation. Differential gene expression analyses were performed using DESeq2 (v1.48.1), correcting for sequencing batch effects. Raw FASTQ reads were processed via GenPipes v6.0.0, and gene-level quantification was done using kallisto v0.50.0 with the GRCh38 Ensembl v90 annotation. (A) Transcription Factors: HNF4A, PDX1, MAFB, HNF1B. (B) Insulin Secretion and Exocytosis: ABCC8, IAPP, SNAP25, STX1A, VAMP2, KCNJ11, EXOC7, CACNA1D. (C) Beta Cell Maturation: PDX1, IAPP, SLC2A2, ABCC8, KCNJ11, CACNA1D. (D) ECM and Signaling: ADAM22, COL5A1. (E) Glucose Transport and Metabolism: SLC2A2, ALDOB. (F) Post-transcriptional Regulation: A1CF. Each histogram corresponds to gene-level comparisons across relevant genotypes (WT, HNF1A–/–, HNF1A+/–, A251T). Genes shown met an adjusted p-value threshold of <0.05, unless otherwise indicated.
